# Daily associations between outdoor air pollution and sleep quality

**DOI:** 10.64898/2026.09.15.26363130

**Authors:** Carolina Temporão, Pauline Caille, Guillaume Chevance, Lilla Kovács, Róbert Pierson-Bartel, Sára Wolf, Peter Przemyslaw Ujma, Paquito Bernard

## Abstract

**Background:** Growing evidence links ambient air pollution, including fine particulate matter (PM_2.5_), nitrogen dioxide (NO_2_), and ozone (O_3_), to adverse effects on sleep. However, most prior research has relied on chronic exposure metrics and subjective sleep assessments, obscuring short-term dynamics and limiting physiological precision. To address these gaps, we examined associations of same-day and previous-day pollutant levels with both self-reported and device-measured sleep, using an ecologically valid design.

**Methods:** Data were derived from the Budapest Sleep, Traits and Experiences Study, a 7-day observational study in which participants reported their daily activities and sleep quality ratings, and recorded EEG during sleep. Device-measured sleep outcomes included sleep efficiency, %REM sleep, sleep onset latency, and total sleep time. Outdoor concentrations of PM_2.5_, NO_2_, and O_3_ were aggregated from monitoring stations during an individual’s awake period. Linear mixed-effects models with a random intercept for participant were employed to evaluate associations between each pollutant and sleep outcome, being adjusted for age, sex, body mass index, education, weekday, season, temperature, physical activity, screen exposure, and traveling.

**Results:** The analysis revealed that elevated O_3_ exposure on day 1 was associated with lower sleep efficiency and poorer perceived sleep quality on night 2. These findings survived correction for multiple comparisons, and held across multi-pollutant models and sensitivity analyses accounting for depression, insomnia, and chronotype. Notably, no robust day-to-night associations emerged for any pollutant.

**Conclusions:** Overall, this study demonstrates a short-term relationship between outdoor ozone concentration and reduced sleep quality, even at levels deemed safe by current standards, which highlights the need for continued vigilance and regulations to protect health.

## Introduction

Sleep is a multidimensional construct, encompassing satisfaction, efficiency, alertness, timing, and duration (Buysse, 2014), and is essential to both physical and mental well-being (Ramar et al., 2021). Conversely, chronic sleep deprivation significantly raises the risk of premature death, metabolic and cardiovascular diseases (such as heart disease, obesity, and diabetes), stress- and mood-related disorders, and reduced cognitive function in the general adult population (Cappuccio et al., 2010; Chattu et al., 2019; Troxel et al., 2017). Despite these severe implications, sleep deprivation continues to grow as a public health epidemic (Owens et al., 2014), making it increasingly necessary to identify the factors that impair sleep.

While several determinants of sleep, including sociodemographic and lifestyle factors, have already been identified (Billings et al., 2020), environmental factors have increasingly been investigated. Among them, chronic exposure to ambient air pollutants – including fine particulate matter (PM_2.5_), nitrogen dioxide (NO_2_), and ozone (O_3_) – has been associated to adverse sleep outcomes (Cao et al., 2021; Chen et al., 2019; Liu et al., 2020; M. Wang et al., 2025). Notably, air pollution also exhibits substantial day-to-day variability, driven by road traffic, industrial processes, and climatic conditions, which has recently led to a growing interest on pollution-sleep associations over shorter time scales, such as day-to-night associations (i.e., daytime exposure versus nighttime sleep) or delayed associations (e.g., day 1 exposure with night 2 sleep).

From the studies that have addressed these short-term dynamics, exposure to air pollutants on day 1 has been linked to multiple sleep disruptions on night 1 (D1-N1) and/or night 2 (D1-N2): reduced self-reported sleep quality associated with NO_2_ (Gignac et al., 2022); longer yet lighter sleep linked to PM_2.5_ and NO_2_, and shorter sleep linked to O_3_ (P. Zhou et al., 2023); increased sleeplessness tied to PM_2.5_ (Heyes & Zhu, 2019); and a rise in apnea–hypopnea index during sleep associated with O_3_ (Weinreich et al., 2015). However, the generalizability and precision of pollution-sleep associations – both for a fraction of these short-term studies and those on longer timescales – are frequently questioned due to a heavy reliance on self-reported data. Recently, objective sleep-scoring systems have become accessible in ecological settings, offering the physiological precision necessary to bridge this gap. Without a complete understanding of these short-term dynamics, policymakers risk missing critical opportunities for timely and effective intervention.

To address this, the present study evaluated the D1-N1 and D1-N2 associations between key air pollutants (PM_2.5_, NO_2_, and O_3_) and specific sleep measures, including both self-reported sleep quality and device-measured metrics (sleep efficiency, percentage of REM sleep, sleep onset latency, and total sleep time). Through this approach, we aimed to provide a clearer picture of how short-term air pollution exposure influences sleep health.

## Methods

### Study Design and Participant Data

This study leveraged data from the Budapest Sleep, Traits and Experiences Study (BSETS), a multidimensional, multi-day observational study examining sleep and daily activities using an ecologically valid design. The full protocol and available data have been documented separately (Taji et al., 2023). An overview of the BSETS study design, including participant-derived information that is relevant for the current study, is depicted in **Figure 1A**. BSETS received approval from both the Semmelweis University Institutional Review Board (IRB) and the Hungarian Medical Council (Reference No. 7040-7/2021/EÜIG), confirming its adherence to the most recent Declaration of Helsinki. Furthermore, every participant provided written informed consent using an IRB-authorized form, and data collection occurred between May 2021 and April 2023.

**Figure 1.**
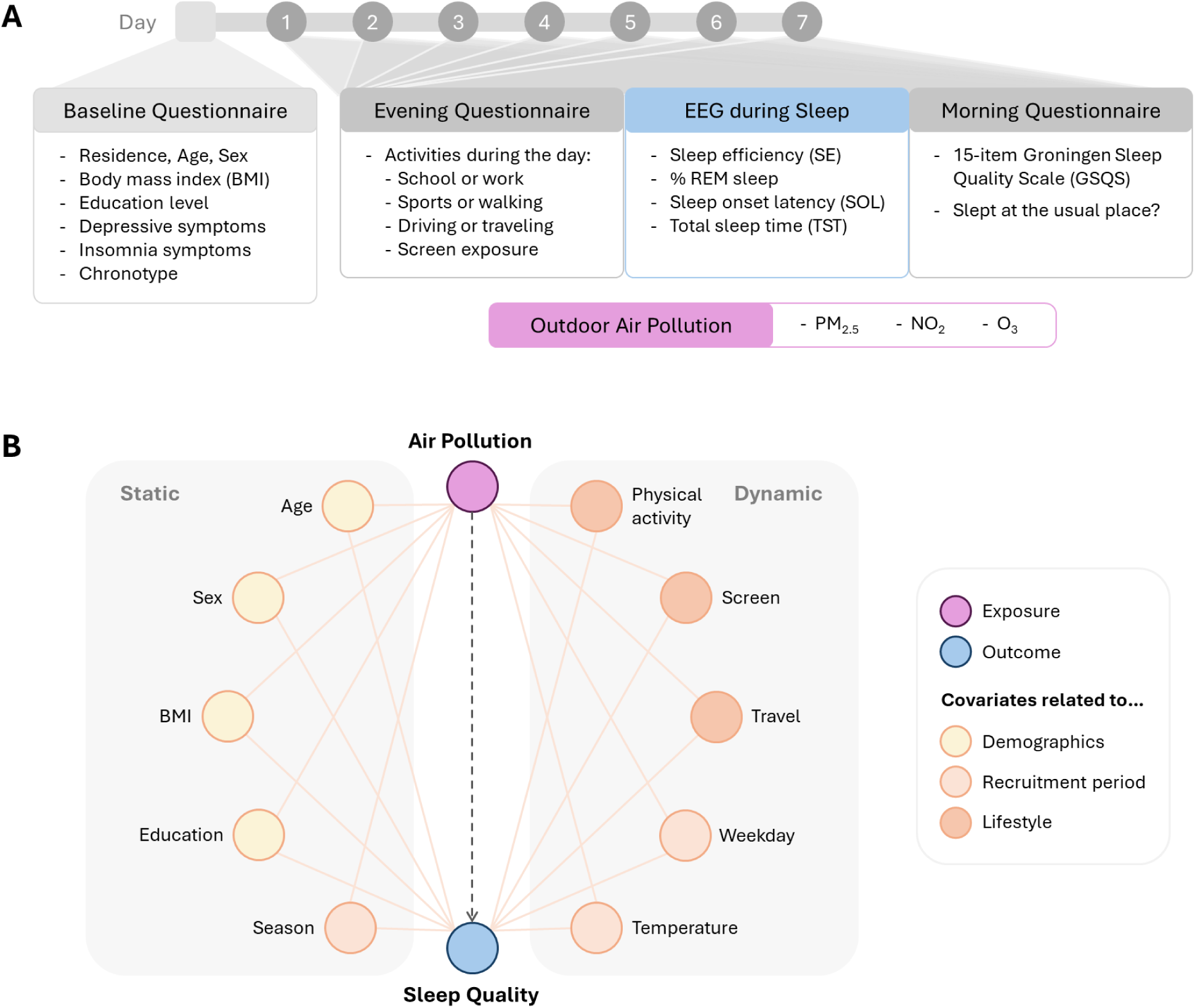
Participant data collection and covariate selection. (A) Participant data was obtained from the Budapest Sleep, Experiences and Traits Study (BSETS), namely demographic and psychological information prior to the study, and lifestyle- and sleep-related data across the seven consecutive days of the study. (B) Directed acyclic graph (DAG) used for covariate selection.

For the initial characterization of the sample, participants completed an extensive questionnaire prior to the study period gathering psychological, demographic, and anthropometric information. Demographic variables included place of residence, age, sex, and education level, along with student status and institution. Height and mass measurements were also obtained to calculate body mass index (BMI). Depressive symptoms, insomnia, and chronotype were also measured using validated questionnaires, respectively: Patient Health Questionnaire (PHQ-9; Kroenke et al., 2001), Athens Insomnia Scale (AIS; Soldatos et al., 2000), and Munich Chronotype Questionnaire (MCTQ; Roenneberg et al., 2003).

Over a seven-day period, participants underwent ecological momentary assessment (EMA) by completing morning and evening questionnaires where, respectively, their sleep quality was rated based on the Groningen Sleep Quality Scale (GSQS; with higher total scores indicating poorer sleep) and their daily activities detailed (see **Figure 1A** and **Supplementary Methods**). Daily activities included going to a workplace or school, practicing sports, walking, traveling or driving for at least one hour, and computer or smartphone engagement. Concurrently, participants wore the validated Dreem2 mobile headband each night to record quantitative electroencephalography (EEG), from which objective sleep metrics derived, as detailed below.

While the BSETS dataset has previously been utilized to examine how internal factors, such as perceived sleep quality, social experiences, emotional involvement, and physical activity, influence sleep quality and variability (Pierson-Bartel & Ujma, 2024; Ujma & Bódizs, 2024, 2026; Wolf et al., 2026), the present study shifts focus to the relationship between air pollution exposure, a prominent external determinant, and sleep quality.

### Eligibility Criteria

The original BSETS dataset comprised 267 participants and 1901 day-level observations, but data from 9 subjects (65 observations) could not be used because the date was missing or duplicated. To ensure meaningful exposure to Budapest air pollution, participants were required to either (1) reside in Budapest and sleep at home on the monitoring nights, or (2) attend classes in the city. Following the application of this criterion and the exclusion of one participant with no information on sleep onset time, the final analytic sample consisted of 190 participants and 1105 observations (1074 for self-reported sleep and 1057 for device-measured sleep).

### Device-measured Sleep Metrics

EEG recordings were automatically scored using an algorithm that demonstrated high validity when compared to visual scoring of gold-standard polysomnography according to American Academy of Sleep Medicine guidelines (Arnal et al., 2020). Objective sleep ratings were extracted from the resulting hypnogram and included total sleep time (TST), sleep efficiency (SE, calculated as the percentage of TST relative to total time in bed, defined as minutes from lights-out to lights-on), sleep onset latency (SOL, measured as minutes from lights-out to the first three consecutive epochs of any sleep stage), and percentage of rapid-eye movement sleep (% REM sleep, representing the percentage of TST spent in REM sleep stage).

### Outdoor Air Pollution

Concentration data for outdoor PM_2.5_, NO_2_, and O_3_ during the study period were sourced from the Hungarian Air Quality Monitoring Network (*Levegőminőség*, 2026). Data were gathered from eight primary monitoring stations located throughout Budapest: Gilice Square, Kőrakás Park, Széna Square, Teleki Square, Budatétény, Erzsébet Square, Gergely Street, and Honvéd. The Pesthidegkút station was excluded from this dataset, consistent with methodologies in prior studies (Mészáros et al., 2025), because it is situated in a hilly region of Budapest and exhibits pollution impacts that differ significantly from those observed at urban stations. To derive representative concentration values, the levels of each air component were averaged across the eight stations, leveraging their high correlations. An exception was made for O_3_ concentrations, which were not collected at the Erzsébet Square and Honvéd stations; for these measurements, averages were computed using data from the remaining six stations.

Exposure was defined individually for each participant and day, using a dynamic window beginning two hours after wake time and ending two hours before sleep onset. When device-measured sleep onset was unavailable, self-reported sleep onset was used as a fallback. In instances where sleep was not monitored the previous night and awakening time was consequently missing, the average awakening time across the entire study period for that specific participant was utilized as a substitute. The same extrapolation approach was applied to sleep onset time. Both same-day and previous-day air pollution levels relative to nighttime sleep were incorporated into the analysis to independently capture immediate (day 1 exposure to night 1 sleep; D1-N1) and lagged effects (D1-N2).

For the estimation of inter-station daily heterogeneity in pollutant concentrations, daytime was roughly defined from 9am to 10pm, and a coefficient of variation was calculated per day and pollutant, as the standard deviation divided by the average of values derived from all available stations.

### Statistical Analyses

To examine associations between daytime outdoor air pollutant levels and sleep quality metrics, we employed linear mixed-effects models using the *lme4* package in R statistical software (Version 4.5.1). By incorporating a random intercept at the individual level, this approach accounted for the nested structure of the data and controlled for within-subject correlation in sleep outcomes across days. More details about this statistical strategy are available in the **Supplementary Methods**.

In separate linear mixed models, we evaluated associations of each pollutant (PM_2.5_, NO_2_, O_3_) with each sleep outcome (GSQS, SE, %REM, SOL, TST), resulting in a total of 5 tests per exposure per timepoint (D1-N1 or D1-N2) and 30 tests globally. Since air pollutants often co-occur and share common sources, we also conducted multi-pollutant models, incorporating PM_2.5_, NO_2_, and O_3_ simultaneously, resulting in additional 10 tests.

All models controlled for potential confounding variables selected based on their documented association with both air pollution exposure and sleep health (see **Supplementary Methods**). As depicted in the directed acyclic graph (DAG) in **Figure 1B**, these covariates relate to demographic or anthropometric factors, external factors associated with the recruitment period, or lifestyle factors, and can be further categorized based on their temporal stability into static and dynamic variables. Static covariates included age, sex, education level, body mass index, and season. Dynamic covariates, in turn, included a binary weekday/weekend variable, average ambient temperature, physical activity, and binary variables for long screen time and long traveling, defined as at least one hour of engagement in each activity. Daily average temperature data were collected from the ERA-5-Land dataset (Copernicus Climate Change Service, 2024; accessed on 6/03/2026) across Budapest during the study period.

For each sleep outcome, we used the following model specification, where *i* and *d* denote individual and day, respectively, whereas *p* denotes pollutant and *j* lag day (*j*=0 in D1-N1 analyses and *j*=1 in D1-N2 analyses):

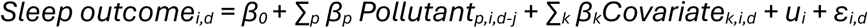

Note that in single-pollutant models, only one pollutant term was included. All models included a random intercept for subject (*u_i_*) and a residual error term (*ε_i,d_*), and were adjusted for the same set of covariates:

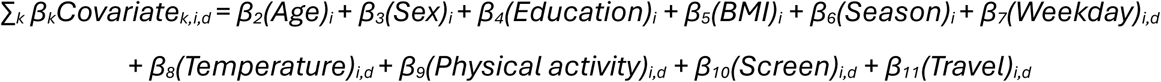

### Sensitivity Analyses

Sensitivity analyses were conducted to determine whether the observed associations between air pollution and sleep quality varied in magnitude or direction depending on specific psychological and sleep-related characteristics: depressive symptoms (PHQ-9 total score), insomnia symptoms (AIS total score), and chronotype (total score of MCTQ-derived midsleep time on free days corrected for oversleeping, MCTQ-MSFsc). To address this, separate multilevel models were run for each sensitive variable by adding it as an additional covariate to the full model. They were computed as continuous parameters, given their distributions and clinically significant thresholds, as shown in **Supplementary Figure 4** and described in the Results section below. Similarly, we controlled for previous-night sleep quality (N0 Sleep in D1-N1 analyses, and N1 Sleep in D1-N2 analyses) in an additional sensitivity analysis for each exposure-outcome model, using the same sleep metric as the outcome.

Statistical significance was set at *p* = 0.05, and we assessed whether significance remained resistant to Benjamini-Hochberg false discovery rate (FDR) correction, which was applied to each family of tests (one per timepoint, each including five outcomes), thus correcting for five tests. Multi-collinearity diagnostics were performed using Variance Inflation Factors (VIF) from the *car* package (Fox et al., 2001), and results are available in **Supplementary Results**.

## Results

### Descriptive Statistics

A total of 190 participants contributed to the analysis of sleep metrics, yielding 1105 day-level observations. **Table 1** shows observation-level statistics for air pollutants, sleep outcomes, and dynamic covariates, alongside subject-level statistics for static covariates and psychological and sleep-related measures used in sensitivity analyses. Variable distributions are also displayed in **Supplementary Figures 1-4**.

**Table 1.** Descriptive statistics.

| Variable | Level | N | Mean (SD) | Median [IQR] | Range | %Missing |
| --- | --- | --- | --- | --- | --- | --- |
| **Air pollutants** |  |  |  |  |  |  |
| PM<sub>2.5</sub> (µg/m<sup>3</sup>) | Observation | 1105 | 14.8 (8.0) | 13.8 [8.5, 19.6] | 1.9 - 48.9 | 0 |
| NO<sub>2</sub> (µg/m<sup>3</sup>) | Observation | 1105 | 29.3 (10.7) | 28.0 [21.4, 35.6] | 6.8 - 63.8 | 0 |
| O<sub>3</sub> (µg/m<sup>3</sup>) | Observation | 1105 | 44.4 (24.2) | 42.0 [23.9, 65.1] | 4.2 - 124.7 | 0 |
| **Sleep outcomes** |  |  |  |  |  |  |
| GSQS score | Observation | 1074 | 4.0 (3.3) | 3.0 [1.0, 6.0] | 0.0 - 14.0 | 2.8 |
| Sleep efficiency (%) | Observation | 1057 | 91.5 (5.8) | 93.0 [89.4, 95.2] | 50.0 - 98.7 | 4.3 |
| REM sleep (%) | Observation | 1057 | 26.1 (7.1) | 25.9 [21.8, 30.1] | 0.0 - 62.2 | 4.3 |
| Sleep onset latency (min) | Observation | 1057 | 14.4 (13.8) | 10.5 [6.0, 17.5] | 1.0 - 152.5 | 4.3 |
| Total sleep time (min) | Observation | 1057 | 401.1 (87.4) | 405.0 [354.0, 458.0] | 50.5 - 671.0 | 4.3 |
| **Static covariates** |  |  |  |  |  |  |
| Sex | Subject | 190 |  |  |  | 0 |
| Female |  | 103 (54.2%) |  |  |  |  |
| Male |  | 87 (45.8%) |  |  |  |  |
| Age (years) | Subject | 190 | 27.6 (11.7) | 22.0 [20.0, 31.0] | 18.0 - 76.0 | 0 |
| BMI (kg/m<sup>2</sup>) | Subject | 185 | 22.9 (3.5) | 22.3 [20.3, 24.7] | 15.4 - 33.2 | 2.6 |
| Education level | Subject | 190 |  |  |  | 0 |
| Secondary |  | 111 (58.4%) |  |  |  |  |
| Bachelor |  | 42 (22.1%) |  |  |  |  |
| Master |  | 32 (16.8%) |  |  |  |  |
| Elementary |  | 5 (2.6%) |  |  |  |  |
| Season | Subject | 190 |  |  |  | 0 |
| Autumn |  | 87 (45.8%) |  |  |  |  |
| Winter |  | 51 (26.8%) |  |  |  |  |
| Spring |  | 42 (22.1%) |  |  |  |  |
| Summer |  | 10 (5.3%) |  |  |  |  |
| **Dynamic covariates** |  |  |  |  |  |  |
| Weekday | Observation | 1105 |  |  |  | 0 |
| Weekday |  | 830 (75.1%) |  |  |  |  |
| Weekend |  | 275 (24.9%) |  |  |  |  |
| Mean temperature (°C) | Observation | 1105 | 7.9 (6.0) | 7.3 [3.6, 11.8] | -3.8 - 30.1 | 0 |
| Physical activity (min) | Observation | 865 | 79.3 (84.1) | 60.0 [30.0, 105.0] | 0.0 - 770.0 | 21.7 |
| Screen time | Observation | 1010 |  |  |  | 8.6 |
| ≥ 1 hour |  | 837 (82.9%) |  |  |  |  |
| < 1 hour |  | 173 (17.1%) |  |  |  |  |
| Drive/travel | Observation | 1102 |  |  |  | 0.3 |
| ≥ 1 hour |  | 585 (53.1%) |  |  |  |  |
| < 1 hour |  | 517 (46.9%) |  |  |  |  |
| **Psychological and sleep-related features** |  |  |  |  |  |  |
| Depression (PHQ-9) | Subject | 185 | 6.1 (4.7) | 5.0 [3.0, 8.0] | 0.0 - 24.0 | 2.6 |
| Insomnia (AIS) | Subject | 188 | 4.5 (3.2) | 4.0 [2.0, 6.0] | 0.0 - 14.0 | 1.1 |
| Chronotype (MCTQ-MSFsc) | Subject | 116 | 4.4 (1.1) | 4.4 [3.6, 5.1] | 2.1 - 7.4 | 38.9 |

The study population comprised predominantly healthy young adults with a median age of 22 years and an average BMI of 22.9, of whom 54.2% were female. Clinical screening indicated that depressive and insomnia symptoms were generally mild and did not reach moderate or extreme clinical thresholds for the majority of the cohort. Specifically, 81.1% of participants exhibited no or mild depressive symptoms, defined as a PHQ-9 score of 9 or lower (Kroenke et al., 2001). Regarding insomnia, the mean score on the AIS test was 4.5 (SD = 3.2). While 23.4% of participants scored above the recommended cutoff of 6 points, only 10.6% reported moderate to severe insomnia severity (defined as an AIS score of 10 or higher) (Soldatos et al., 2000). Given these distributions, variables used in sensitivity analyses, including chronotype, were treated as continuous parameters. Chronotype assessment revealed that 70.7% of participants did not fall into distinct early or late categories (MCTQ-MSFsc scores < 3.5 or > 6.5, respectively) (Roenneberg et al., 2003).

Regarding covariates distribution, it is important to note that summer months were significantly underrepresented, accounting for only 5.3% of the observations, and that information regarding physical activity duration was missing for 21.7% of the sample. Restricting attention to observations with complete data across exposure, outcome, and covariates yields an analytical sample of 147 participants (704 observations) for associations between air pollution and self-reported sleep quality and 143 participants (682 observations) for associations with device-measured metrics.

### Air Pollution Variability

In this study, daily air pollutant concentrations were calculated as the arithmetic mean across all urban monitoring stations in Budapest. To assess the reliability of these aggregated estimates, we quantified the relative spatial heterogeneity of pollutant concentrations across stations for each day using the coefficient of variation (CV). On average, monitoring stations disagreed by 34% for PM_2.5_ concentrations, 36% for NO_2_, and 28% for O_3_ (**Supplementary Figure 1B**).

To visualize exposure variance across the sample, we plotted a time-course of average day concentrations for each pollutant, defined based on a dynamic, subject-specific timeframe (**Figure 2A**). These analyses revealed that the percentage of days where air quality was not fair, according to European Environment Agency guidelines (EEA; González Ortiz et al., 2025), was as follows: 44.4% for PM_2.5_ (>15 μg/m^3^); 61.5% for NO_2_ (>25 μg/m^3^); 0.9% for O_3_ (>100 μg/m^3^).

**Figure 2.**
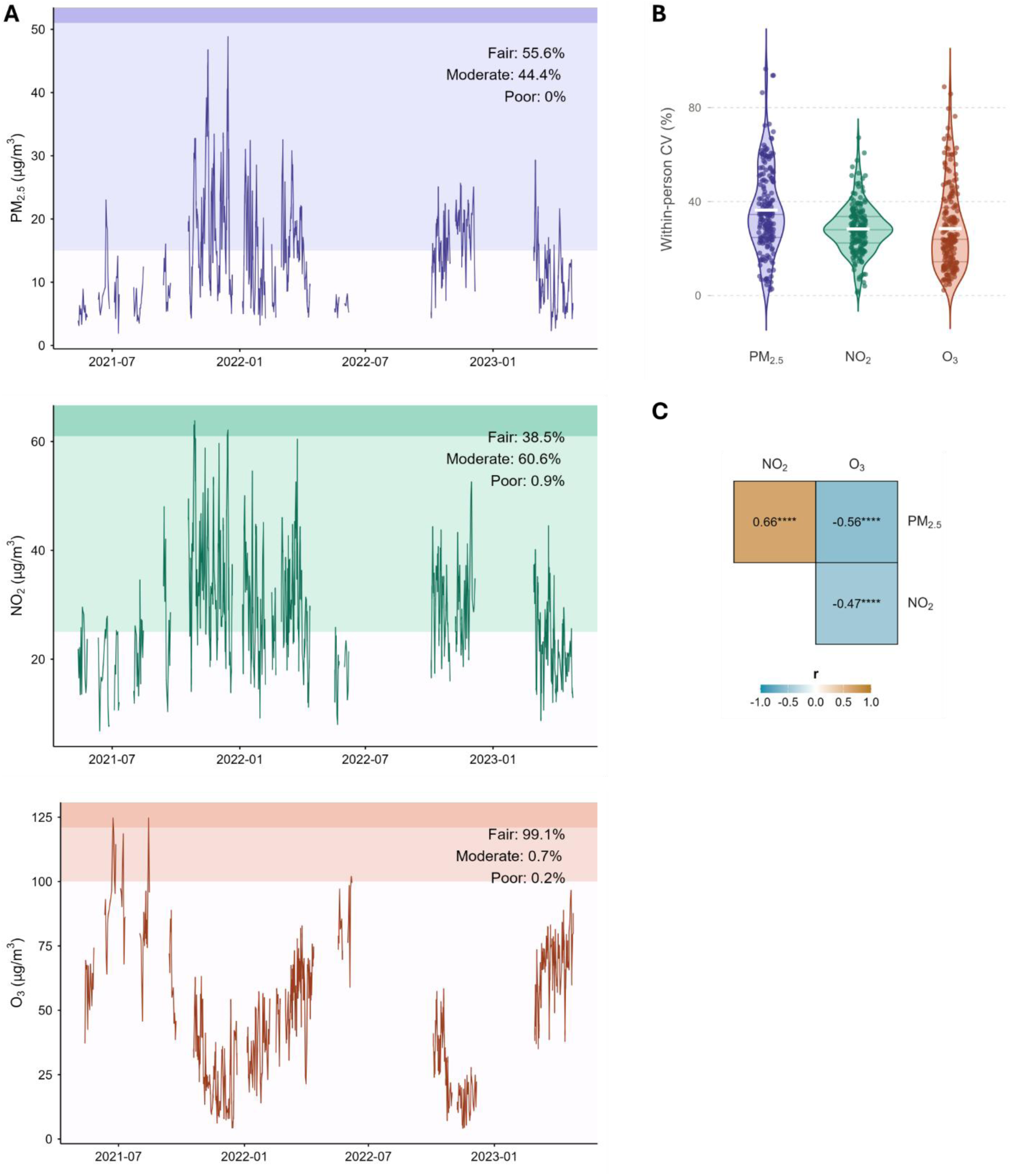
Air pollution variability. (A) Time-series of air pollutant concentrations, defined based on a dynamic, subject-specific timeframe; periods absent from the study were roughly masked (with a minimum gap of 5 days), and distinct background colors represent fair (lightest), moderate and poor (darkest) air quality as defined by EEA guidelines. (B) Within-subject coefficient of variation for air pollutant concentrations across days; the violin plots display individual datapoints representing each participant’s CV, as well as average (white bar) and quartile lines. (C) Pearson’s correlations across air pollutant concentrations. **** p < 0.0001.

We observed considerable day-to-day variability in personal pollutant exposure among participants (**Figure 2B**). While some individuals experienced stable air quality (CV as low as 1.3%), others showed substantial fluctuations (CV up to 96%), with an average within-person coefficient of variation of 36% for PM_2.5_, and 28% for NO_2_ and O_3_. This variability encouraged us to examine how daily fluctuations in air pollution relate to sleep; in other words, whether there are same-day effects and/or lagged effects of outdoor pollution exposure on sleep.

We then computed correlations among air pollutant levels and found all pairwise correlations to be statistically significant (**Figure 2C**). Whereas PM_2.5_ and NO_2_ levels were positively correlated (Pearson’s r = 0.66), O_3_ showed negative correlations with each of these pollutants (Pearson’s r = −0.47 with NO_2_ and r = −0.56 with PM_2.5_). Considering these values, our analyses employed both single- and multi-pollutant models.

### D1-N1 Associations Between Air Pollution and Sleep

To investigate D1-N1 associations between air pollution exposure and sleep outcomes, we employed linear mixed-effects models with random intercepts by participant, controlling for both static and dynamic covariates. In addition to the main models, we conducted multiple sensitivity analyses accounting for depression, insomnia, chronotype, and previous-day (N0) sleep (**Figure 3**). Comprehensive statistical summaries are provided in **Supplementary Tables 1-3**.

**Figure 3.**
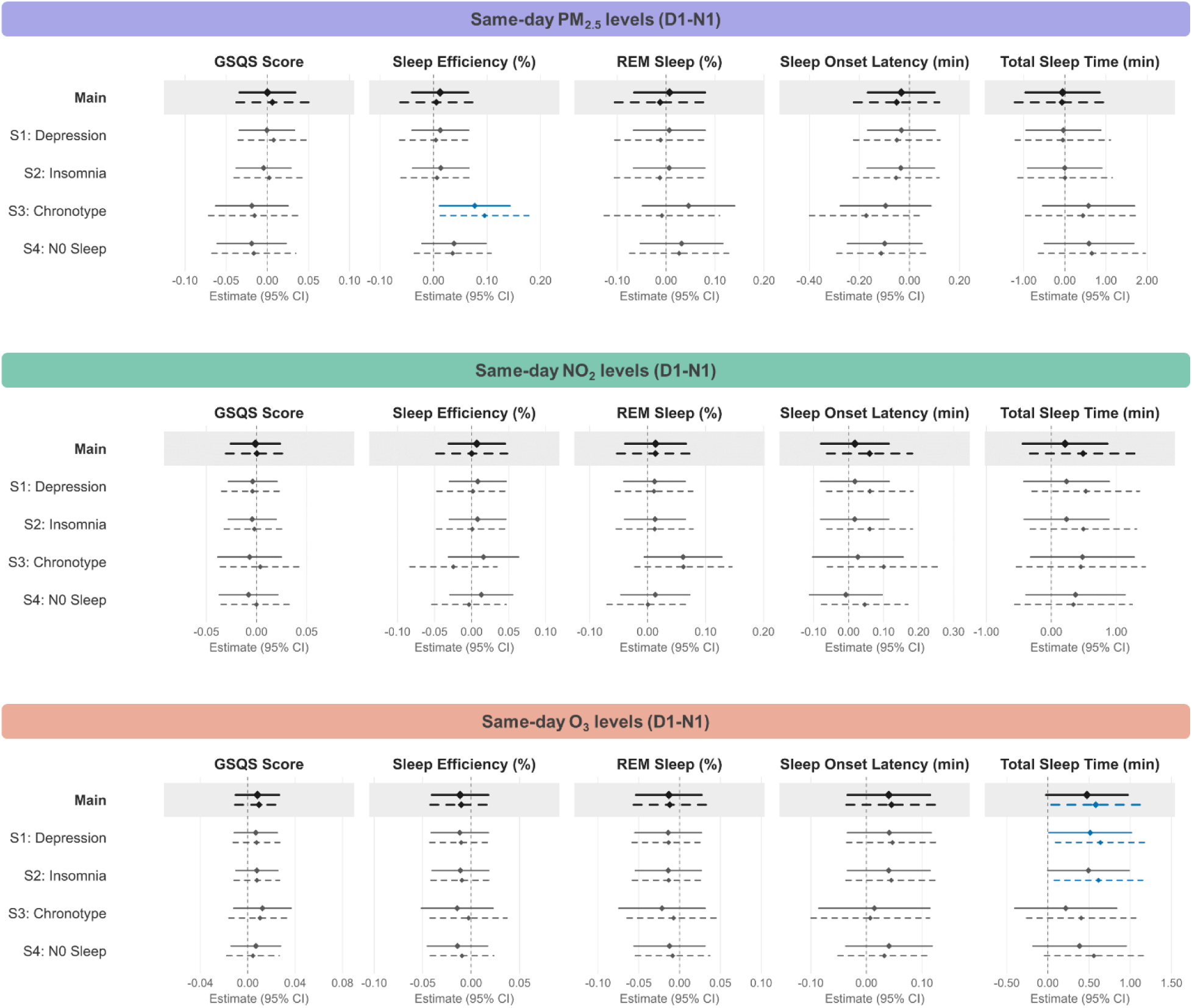
Estimates (95% CI) related to day-to-night (D1-N1) associations between daytime levels of PM_2.5_ (top), NO_2_ (middle), and O_3_ (bottom) and multiple sleep outcomes. Results from the main models (grey background) are followed by results from multiple sensitivity analyses, adjusting for scores of depression (S1), insomnia (S2), or chronotype (S3), or for previous-night (N0) sleep (S4), in addition to covariates. Solid lines represent single-pollutant models, whereas dashed lines represent multi-pollutant models. Blue color indicates p < 0.05 before, but not after, FDR correction.

Overall, no robust D1-N1 associations between pollution and sleep emerged. For PM_2.5_, in contrast to the main models, a sensitivity analysis adjusting for chronotype revealed a positive association with sleep efficiency in both single- and multi-pollutant models. Conversely, higher daytime O_3_ exposure was associated with longer sleep at night, both in the main model and when controlling for depression or insomnia scores, but these effects were restricted to the multi-pollutant approach. Estimates suggested an average increase of 24 minutes in total sleep time for every interquartile range (IQR) µg/m^3^ rise in daytime outdoor O_3_ concentration (equivalent to 41.1 µg/m^3^). None of these associations, however, resisted correction for multiple testing.

### D1-N2 Associations Between Air Pollution and Sleep

Given the potential for delayed physiological responses to environmental stressors, we next examined lagged effects, specifically the association between exposure on day 1 and sleep quality on night 2 (D1-N2 associations; **Figure 4**). Full statistical summaries are available in **Supplementary Tables 4-6**.

**Figure 4.**
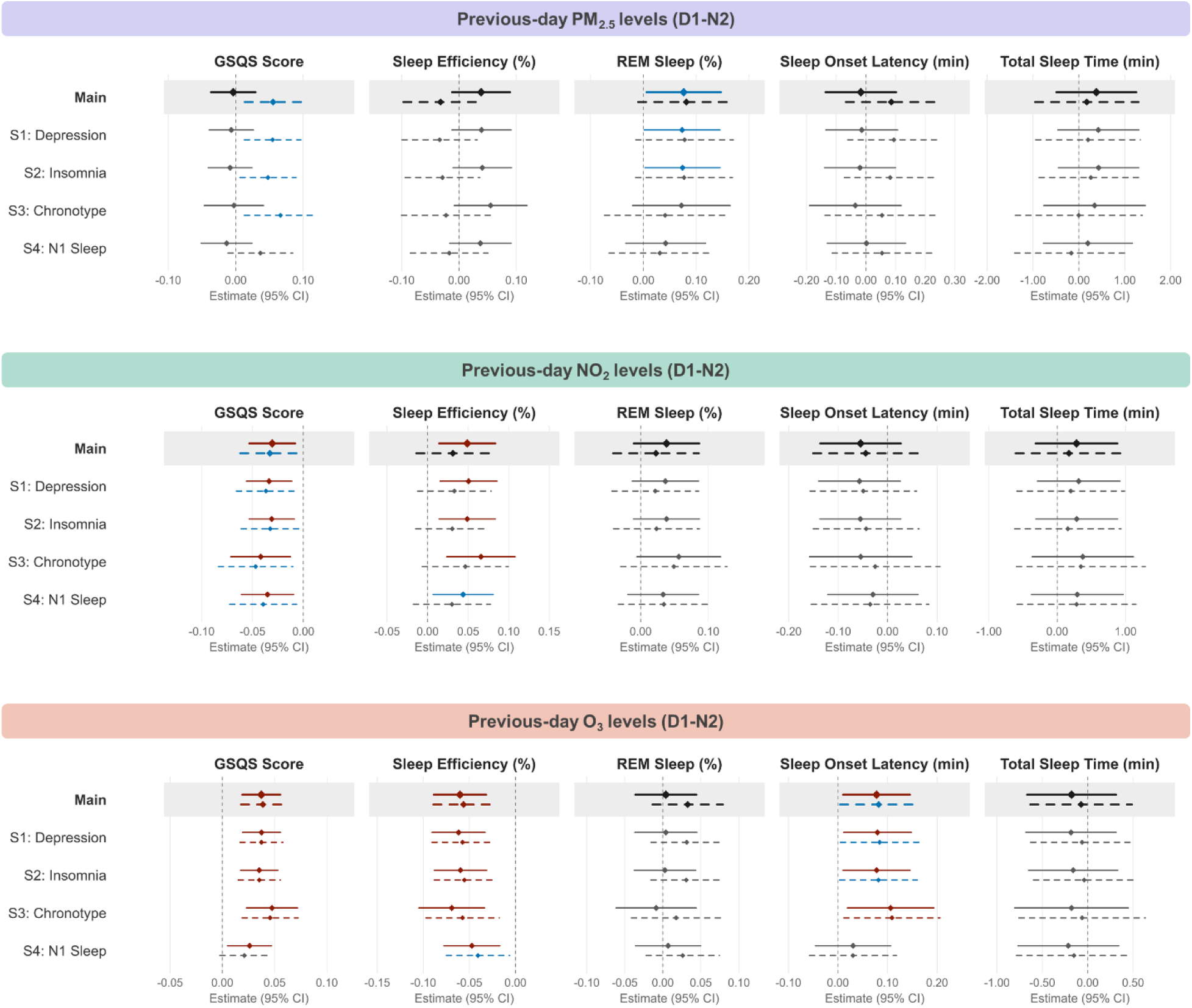
Estimates (95% CI) related to lagged (D1-N2) associations between day levels of PM_2.5_ (top), NO_2_ (middle), and O_3_ (bottom) and multiple sleep outcomes. Results from the main models (grey background) are followed by results from multiple sensitivity analyses, adjusting for scores of depression (S1), insomnia (S2), or chronotype (S3), or for previous-night (N1) sleep (S4), in addition to covariates. Solid lines represent single-pollutant models, whereas dashed lines represent multi-pollutant models. Red color indicates p < 0.05 both before and after FDR correction, whereas blue color indicates p < 0.05 only before correction.

In the single-pollutant models, PM_2.5_ showed no D1-N2 associations with sleep, aside from an FDR-sensitive link to %REM sleep. NO_2_ and O_3_, by contrast, showed robust (FDR-resistant) associations with both GSQS and sleep efficiency across the main models and nearly all sensitivity analyses, though in opposite directions. Each IQR µg/m^3^ increase in outdoor O_3_ on day 1 was associated with an average 1.53-point increase in GSQS (indicating poorer sleep) and a 2.48% decrease in sleep efficiency on night 2, whereas every IQR µg/m^3^ increase in NO_2_ (equivalent to 14.2 µg/m^3^) was associated with a 0.43-point decrease in GSQS and a 0.69% increase in sleep efficiency.

Since these opposing associations mirror the inverse relationship between NO_2_ and O_3_ levels (**Figure 2C**), we next examined whether they held in multi-pollutant models. The associations with GSQS remained FDR-resistant for O_3_, while those for PM_2.5_ and NO_2_ became FDR-sensitive; this pattern held across all models except the sensitivity analyses controlling for N1 GSQS, where associations for PM_2.5_ and O_3_ were non-significant. Similarly, the O_3_-sleep efficiency association remained FDR-resistant across all models except when controlling for N1 GSQS, while the corresponding associations for PM_2.5_ and NO_2_ were non-significant throughout. Overall, O_3_ was the only pollutant with consistently FDR-resistant D1-N2 associations with both GSQS and sleep efficiency, regardless of the number of pollutants included in the model, and these associations persisted after adjusting for depression, insomnia, and chronotype.

Beyond its associations with GSQS and sleep efficiency, O_3_ was also robustly associated with sleep onset latency across all single-pollutant models except the sensitivity analysis adjusting for N1 sleep. Main model estimates indicated a 3.2-minute increase in sleep onset latency on night 2 for every IQR µg/m^3^ rise in O_3_ on day 1. In the multi-pollutant models, however, this association remained significant only in the chronotype-adjusted model regardless of multiple testing correction.

To begin exploring mechanisms underlying these lagged effects, we adjusted D1-N2 models for day 2 awake time, which may influence homeostatic sleep drive. After this adjustment, only the associations between O_3_ and GSQS and between O_3_ and sleep efficiency remained statistically significant, both before and after FDR correction, across single- and multi-pollutant models (**Supplementary Table 6**).

## Discussion

The main objective of this study was to elucidate short-term associations between air pollution exposure and sleep outcomes in an urban population. Using an ecological approach that accounted for a relevant range of demographic, behavioral, and environmental factors, we found that pollution-sleep associations differed markedly according to the temporal scale of exposure (D1-N1 versus D1-N2), the pollutant considered, and the adjustment strategies employed.

The most robust finding to emerge from this study concerns O_3_ exposure, which showed a negative D1-N2 association with both self-reported sleep quality and device-measured sleep efficiency. This association held in both single- and multi-pollutant models, resisted correction for multiple testing, and persisted across sensitivity analyses, which ruled out the confounding effect of depression, insomnia, and chronotype. These results align with a growing body of literature linking short-term O_3_ exposure to poorer sleep health, including associations with sleep-disordered breathing (Weinreich et al., 2015) and with increased hospital visits for sleep disorders (Tang et al., 2020). Long-term exposure has also been linked to higher rates of wheezing-related sleep disturbance (Sánchez et al., 2019) and worse self-reported sleep quality (Kheirandish-Gozal et al., 2014) in children, and 3-year averaged O_3_ exposure has been associated with increased insomnia symptoms (Xu et al., 2021). More directly comparable to the present study, Zhang and colleagues (2023) examined lagged effects of day-level O_3_ exposure on accelerometer-measured sleep architecture. They found significant associations between cumulative lag-1 O_3_ exposure and both sleep efficiency and sleep onset latency, but not total sleep time (Y. Zhang et al., 2023), which closely mirrors our single-pollutant D1-N2 results.

Several biological mechanisms may plausibly underlie the observed relationship between O_3_ exposure and sleep (Cao et al., 2021; Nuvolone et al., 2018). Perhaps the most compelling involves ozone-mediated neurotransmitter imbalance (for example, via the dysregulation of serotonin metabolism) in brain regions central to sleep-wake regulation, contributing to both nocturnal insomnia and daytime sleepiness (Alfaro-Rodríguez & González-Piña, 2005; Bertini et al., 2010; Portas et al., 2000). Beyond this, O_3_ exposure appears to promote neuroinflammation and neurodegenerative processes within the central nervous system, potentially impairing the neural circuitry that governs sleep (Araneda et al., 2008; Brockmeyer & D’Angiulli, 2016; González-Guevara et al., 2014). A third pathway involves the respiratory system: O_3_ causes respiratory cell injury and triggers downstream inflammatory cascades and neuroendocrine stress responses (Bush et al., 1996; Sarangapani et al., 2003), which may restrict airflow and heighten the risk of sleep-disordered breathing (Weinreich et al., 2015). Determining which of these mechanisms, or combination thereof, drives the effects observed here, and why they emerge with a lag rather than immediately, will require further study. Notably, our sensitivity analyses ruled out reduced homeostatic sleep drive as an explanation for the lagged effects of O_3_ on sleep.

In terms of effect size, our model estimates indicate that each IQR increase in outdoor O_3_ on day 1 was associated with an average 1.53-point increase in GSQS and a 2.48% decrease in sleep efficiency on night 2. Good sleep in (young) adults is typically defined as sleep efficiency above 85%, a threshold met by 91% of day-level observations in our sample. On the surface, this might suggest that ozone’s impact on sleep, though statistically significant, is not practically meaningful in this population. We argue, however, that this impact should not be underestimated, for several reasons. First, the shift in sleep physiology was pronounced enough to be consciously perceptible, as reflected in participants’ self-reported worsening of sleep quality. Second, outdoor daytime O_3_ concentrations peak (and frequently exceed health-safe thresholds) in summer months, but these were highly underrepresented in this sample, meaning the true effects during periods of high exposure may be considerably stronger. Third, and relatedly, climate change is expected to accelerate the photochemical reactions that generate tropospheric O_3_ (Vazquez Santiago et al., 2024). Without mitigation and adaptation measures, future populations could face both higher O_3_ concentrations and correspondingly greater sleep disruption.

Given the consistency of these effects across analytical strategies, O_3_ stands out as the strongest candidate among the pollutants studied for a genuine, physiologically meaningful effect on sleep. Notably, NO_2_ also showed robust associations with both self-reported sleep quality and sleep efficiency in single-pollutant models, but in the opposite direction, i.e., associated with better sleep outcomes. This divergence likely reflects the atmospheric chemistry linking the two pollutants: O_3_ and NO_2_ covary substantially on a day-to-day basis, but inversely, since NO_2_ typically co-occurs with the ozone precursors (NO_X_) that rapidly deplete ambient ozone (Jaroszyńska-Wolińska, 2010). The fact that associations with self-reported sleep quality and sleep efficiency remained robust for O_3_ but not for NO_2_ in multi-pollutant models suggests that the opposing valence observed for NO_2_ may be an artifact of this inverse atmospheric relationship rather than a genuine protective effect. This interpretation is further supported by the broader literature, which consistently links long-term NO_2_ exposure to poorer self-reported sleep health, including worse sleep duration and habitual sleep efficiency (H. Wang et al., 2024), longer sleep onset latency (Y. Wang, Liu, et al., 2020), and greater daytime sleepiness (Y. Wang, Mao, et al., 2020). Our findings diverge notably from those of Gignac and colleagues (2022), who reported negative daily-level associations between NO_2_ and self-reported sleep quality (Gignac et al., 2022). Reassuringly, despite this inverse relationship between O_3_ and NO_2_, no multicollinearity concerns arose in any of the multi-pollutant models in the present study. Taken together, these results underscore the value of multi-pollutant modeling and the complexity of interactions among air pollutants: combined exposure to multiple pollutants may obscure or reveal sleep-related effects that isolated single-pollutant analyses would otherwise miss.

### Strengths and Limitations

This study distinguishes itself through three pivotal strengths. First, by leveraging mobile EEG technology, we achieved an ecological and fine-grained characterization of sleep that surpasses traditional methods. Second, we ensured statistical rigor and transparency through a dual reporting strategy for multiple comparisons and pollutant modeling; this approach allowed us to stratify findings by robustness, identifying the most trustworthy results as those retaining significance across all analytical strategies. Finally, our investigation benefits from a unique epidemiological profile, with a substantial sample size of over 1000 nights within a demographic of healthy, working-age adults. Conducting this research in Budapest – a medium-large European city – provides critical geographical diversity, addressing a gap where most existing studies concentrate on either highly polluted Asian metropolises or North American contexts, thereby enhancing the generalizability of our findings across varied urban environments.

While this study offers significant insights, several limitations must be acknowledged. First, all participants were assumed to share the same air pollution exposure levels across Budapest on any given day. This is concerning given both the substantial inter-station variability and the findings of Gignac et al. (2022), which demonstrated that associations between NO_2_ and self-reported sleep emerged only when using aggregate station data, disappearing when exposure was matched with individual mobility data or collected from passive diffusion tubes. Methodological constraints also arose in our lagged analyses and relate to the 15% of participants residing in areas bordering Budapest. Although they attended classes within the city and their daytime exposures were likely comparable to residents, it is uncertain whether they were in Budapest the day prior to sleep screening; nevertheless, this concerned only a small proportion of the sample and is unlikely to materially affect the results. Furthermore, the inclusion of dynamic covariates time-matched to sleep outcomes but not with pollution exposure in lagged models may have introduced outcome-biased influence. Finally, endogenous covariates were self-reported, which limited physiological precision. Among them, lifestyle variables were further constrained by binary operationalizations with fixed cut-offs (e.g., screen time and travel time dichotomized at one hour) or captured only by duration without accounting for intensity (as it was the case of physical activity), potentially obscuring nuanced relationships.

### Future Directions

The present findings encourage future research to combine both self-reported and objective approaches for a more comprehensive assessment of sleep, and to employ multi-pollutant modelling for a more integrated assessment of exposure. Future work should also address several critical avenues to deepen our understanding of the pollution-sleep nexus. To begin, it remains an open question whether the observed associations persist under distinct environmental conditions or within more vulnerable populations. Specifically, investigations are needed during summer months, a period where ozone concentrations often reach levels above EEA-defined health thresholds, and where prior studies have notably identified short-term associations between air pollution (specifically PM_10_) and sleep disturbances (Zanobetti et al., 2010). Furthermore, future studies would benefit from a more robust technological characterization of both exposure and lifestyle behaviors. Moving beyond aggregate data, researchers could incorporate geolocation tracking or mobile monitoring devices to capture individual-level pollution exposure with greater precision (Kerckhoffs et al., 2025), while simultaneously integrating theoretically relevant variables such as noise and light pollution, as well as access to green and blue spaces, which may confound the observed associations with sleep (Son et al., 2021; Yasmeen et al., 2025). Finally, there is a distinct need to disentangle the effects of indoor versus outdoor pollution and diurnal versus nighttime exposure on sleep outcomes. Such inquiries would be significantly enriched by measuring behavioral and technological adaptations, including adherence to sleep hygiene practices and the utilization of mitigation strategies like air filters, thereby offering a more comprehensive view of how individuals navigate and respond to polluted environments.

### Concluding Remarks

Sleep is increasingly recognized as critical to individual and societal well-being, particularly amid a growing global sleep deficit (Freeman et al., 2020; Irish et al., 2015; Marjot et al., 2021). Air pollution represents a comparably pressing global health challenge, one intensified by urbanization (X. Zhang et al., 2022), with day-to-day fluctuations frequently exceeding health-based thresholds (*WHO Global Air Quality Guidelines*, 2021). International efforts have made progress in reducing pollutant levels through stricter regulation (European Environment Agency, 2026a; *WHO Global Air Quality Guidelines*, 2021). In Hungary specifically, permit limits, residential energy modernization, improved waste management, and public awareness campaigns have driven substantial gains over recent decades (*Health Impacts of Air Pollution*, 2025), though further progress is still needed to meet national air quality targets.

Current air quality guidelines are built primarily around well-established short- and long-term health outcomes, with sleep rarely, if ever, factored into how these thresholds are set. Yet, our study’s most robust finding – the detrimental association between ozone and sleep quality – emerged even though ozone levels in our sample rarely exceeded moderate thresholds. This suggests that even pollutant concentrations within current guidelines may be sufficient to compromise sleep health, raising the possibility that explicitly incorporating sleep as a health outcome could justify more stringent standards. Protecting sleep would thus function as an upstream preventive strategy, intervening at one of the earliest physiological pathways linking air pollution to long-term health.

## Supporting information

Supplementary Methods; Supplementary Results; Supplementary Figures; Supplementary Tables

## Data Availability

All data produced in the present study are available upon reasonable request to the authors, and our analysis codes are also available on https://doi.org/10.5281/zenodo.21998713.

