## Supplementary Methods; Supplementary Results; Supplementary Figures; Supplementary Tables for "Daily associations between outdoor air pollution and sleep quality"

**Supplementary Materials**

### **Supplementary Methods**

#### Questionnaire-based EMA Data

Over the seven-day study period, participants were asked to complete evening and morning questionnaires daily. In the evening, questionnaires were used to collect information on participant’s daily activities, specifically whether they attended school or work, engaged in sports or walking, drove or traveled beyond walking for at least one hour, or used smart devices for a minimum of one hour. In addition, upon waking each morning, participants provided self-reports regarding the context and setting of their sleep (i.e., usual or unusual location, and alone or accompanied), and sleep quality. The latter was assessed using the Groningen Sleep Quality Scale (GSQS), a 15-item validated scale based on yes/no questions rating sleep quality, with higher total GSQS scores indicating poorer sleep (Pierson-Bartel & Ujma, 2024).

#### Statistical Analyses

Previous BSETS-based studies employed hybrid linear mixed-effects models to decompose within- and between-person effects in associations between day experiences and sleep or between distinct sleep metrics (Pierson-Bartel & Ujma, 2024; Ujma & Bódizs, 2024, 2026; Wolf et al., 2026). While the sample and dependent variables overlapped with the present study, we utilized a naive linear mixed-effects models (LMMs) with random intercept for participant instead. Several reasons were on the basis of such a methodological decision. First, whereas the studies cited above use internal or behavioral factors as independent variables, which may strongly diverge across individuals, we used air pollutant levels, which are purely exogenous. Previous studies examining the relationship between environmental factors, including air pollution and temperature, and EMA-derived internal variables, such as sleep or affect, did not perform within-/between-person decomposition but followed a similar approach to ours, which boosts comparability across studies (Gignac et al., 2022; Montanari et al., 2026; Rocha et al., 2026; P. Zhou et al., 2023). Most importantly, our research question was anchored on the overall association between pollution and sleep, rather than on the within- and between-person effects, which would arguably add unnecessary complexity to the interpretation of findings. Nevertheless, for full disclosure, we calculated intraclass correlation coefficients (ICCs) for each air component and found that the variance of PM_2.5_ and NO_2_ levels was mostly attributable to within-subject differences (ICC[PM_2.5_] = 0.39; ICC[NO_2_] = 0.31), while O_3_ concentration variance had a much stronger between-subject component (ICC = 0.80), which matches with its high seasonality (Ansari et al., 2025).

#### Covariates

The covariates used across all models were selected based on their documented association with both pollution exposure and sleep outcomes, as detailed next.

Static covariates included age as a continuous variable and sex as a binary variable, both standard controls associated with variance in sleep patterns (Rahimi-Eichi et al., 2025) and the effects of short- and long-term air pollution exposure (H. Zhang et al., 2024). Education level was recoded into four standardized categories to serve as a measure of socioeconomic status, a factor known to explain variance in sleep health (Papadopoulos & Sosso, 2023) and exposure impacts (Hajat et al., 2015). Body mass index was also included given evidence of relationships with both air pollution exposure (Huang et al., 2020) and sleep duration (Amiri, 2023; Garfield, 2019). Beyond these demographic and anthropometric covariates, we also considered season, defined as the most represented categorical value in each subject's participation timeframe. Season is a widely-known factor influencing sleep (Scott et al., 2025) and atmospheric circulations affecting air quality (Miao et al., 2015).

Dynamic covariates encompassed factors varying during the recruitment period. First, a binary weekday/weekend variable was included, with Hungarian national holidays classified as weekends, given consistent reports of weekday variations in sleep (Scott et al., 2025) and air quality time series (He, 2023). Moreover, we accounted for ambient temperatures, considering their potential influence on air pollution-sleep associations independently of season. Specifically, higher outdoor temperatures are generally linked to degraded sleep quality (Chevance et al., 2024) and to distinct pollutant concentrations (Kinney, 2008; Steiner et al., 2010), and there is mounting research on synergetic effects of temperature and air pollution on health outcomes (Anenberg et al., 2020; Hu et al., 2022; Orru et al., 2017; Rackow et al., 2025). To account for it, daily average temperature data were collected from the ERA-5-Land dataset (Copernicus Climate Change Service, 2024; accessed on 6/03/2026) across Budapest during the study period.

Finally, lifestyle-related factors were incorporated as dynamic covariates. These included physical activity, derived from self-reported sport and walking time in minutes, and binary variables for long screen time and long traveling or driving, defined as at least one hour of engagement in each activity. Screen exposure, specifically, was coded as 1 only if the participant self-reported at least 60 minutes of watching movies or TV series or reading or playing on a cellphone or another smart device. In the literature, sleep duration and quality were shown to be influenced positively by physical activity (Atoui et al., 2021; Bisson & Lachman, 2023; Dolezal et al., 2017) and negatively by excessive screen time (Deivendran et al., 2025; Hartstein et al., 2024; Zhong et al., 2025), while ambient air pollution is reported to negatively associate with physical activity and positively with leisure-time physical inactivity (An et al., 2018; Kim et al., 2021). Long commuting time was also controlled for as a source of pollutant exposure (Meena & Goswami, 2024; Ramel-Delobel et al., 2024) and a potential cause of suboptimal sleep (Mantripragada & Prasad, 2025).

Prior to fitting the main models, we explored the crude associations between each covariate and sleep outcomes using univariable mixed-effects models, which were compared with intercept-only models using a relaxed significance threshold (α = 0.08). Relative to at least one sleep outcome, such contrast showed associations with all covariates, except sex, season, and traveling (10.5281/zenodo.21998713). Regardless of their crude associations, all covariates identified *a priori* were retained in the adjusted models consistent with a DAG-informed confounding structure, which is arguably more valuable.

#### Sensitivity Analyses

Sensitivity analyses examined whether air pollution–sleep associations varied by participant characteristics: depressive symptoms (PHQ-9), insomnia severity (AIS), chronotype (MCTQ-MSFsc), and prior-night sleep quality (Lag1-Sleep). Below, we detail what justified these analyses.

First, the inclusion of participant’s depressive profile was driven by the well-documented bidirectional relationship between depression and sleep, where sleep disturbances serve as both a primary symptom and a risk factor for depressive disorders (Rosenblum et al., 2025). Furthermore, strong associations have been reported between PM_2.5_ and NO_2_ exposure and depression, alongside moderate links with O_3_, across both short- and long-term periods (European Environment Agency, 2026b).

Second, insomnia is a ubiquitous feature of disordered sleep in both clinical and healthy populations (van Straten et al., 2025), and was included because it can lead to excessive daytime sleepiness, potentially influencing lifestyle choices and, consequently, overall ambient air pollution exposure. Although the specific pathway of insomnia influencing exposure has not been extensively studied, multiple studies confirm the impact of air pollution on insomnia symptoms (Liu et al., 2020; M. Wang et al., 2025).

Third, chronotype is a fundamental determinant of sleep timing and preferences (Yuri et al., 2025). While its direct influence on air pollution exposure levels is less explored, it may affect exposure through variations in the timing of daily activities, such as physical exercise (Vitale & Weydahl, 2017), which can alter pollutant absorption rates (Hahad et al., 2021; Tainio et al., 2021). Conversely, recent evidence suggests that exposure to PM_2.5_ and NO_2_ may itself drive shifts toward later sleep chronotypes in adolescents (Li et al., 2026).

A final sensitivity analysis targeted sleep quality on the previous night (N0 for D1-N1 associations; N1 for D1-N2 associations). Similar to insomnia, prior sleep quality influences both daytime and nighttime sleepiness, which may subsequently modulate an individual's exposure to air pollutants during the subsequent day. By systematically adding these variables, we aimed to verify the robustness of the primary findings against these potential confounding or moderating factors.

### **Supplementary Results**

#### Correlations across sleep outcomes

Although correlations across sleep outcomes have been reported before using the BSETS (Pierson-Bartel & Ujma, 2024), we assessed them for the BSETS subset used here (**Supplementary Figure 2C**). Similar to the previous report, all device-measured metrics were significantly correlated with self-reported sleep (GSQS score), with a positive association appearing for sleep onset latency (Pearson's r = 0.19) and negative associations for the others. Across device-measured metrics, % REM sleep was only significantly correlated with total sleep time (Pearson's r = 0.18), which was, in turn, correlated with all others metrics – positively with sleep efficiency (Pearson's r = 0.32) and negatively with sleep onset latency (Pearson's r = -0.10). Finally, these two sleep outcomes were inversely correlated (Pearson's r = -0.62).

Multicollinearity diagnostics

Multi-collinearity was assessed in each model using the generalized variance inflation factor (GVIF), which takes into account multiple degrees of freedom (df), implemented via the *car* package in R. Following Fox & Monette (1992), the adjusted statistic GVIF^1/(2·df)^ was squared, allowing direct comparison with the standard VIF threshold of 5. No predictor exceeded this threshold in any model (10.5281/zenodo.21998713), indicating acceptable levels of multi-collinearity across all analyses.

Deviations from normality

As shown in **Supplementary Figure 2**, sleep efficiency and sleep onset latency were both highly skewed. As a result, the distributional assumptions underlying LMMs, namely homoskedasticity and normality of residuals, were violated in our analyses of these outcomes.

Prior work using this dataset has addressed this issue in different ways: winsorizing extreme values based on visual inspection (Pierson-Bartel & Ujma, 2024), applying a log-transformation (Ujma & Bódizs, 2026), or fitting generalized, link function-based models to accommodate the skewed distributions (Pierson-Bartel & Ujma, 2024). Winsorizing and log-transformation both have the appeal of relative simplicity, but each carries notable drawbacks. Winsorizing depends on somewhat subjective cutoff for defining extreme values, and discards genuine variability in the data rather than modeling it directly. Log-transformation complicates interpretation, since resulting effects are expressed on the log rather than the original scale, and introduces bias when back-transformed to the raw scale.

To more directly and decisively evaluate the role of non-normality and outliers in our results, we instead fit generalized linear mixed models (GLMMs) using a distribution matched to the shape of each outcome. For sleep efficiency, which is bounded between 0-100% (rescaled to a 0-1 proportion), we used a Beta GLMM with a logit link, which accommodates both the boundedness and the skew of the distribution. For sleep onset latency, which exhibits a floor effect near zero, we used a Gamma GLMM with a log link.

Because GLMM coefficients are estimated on the link scale (log-odds for the Beta/logit model, log units for the Gamma/log model), we additionally computed average marginal effects on the probability scale using the *marginaleffects* package, which back-transforms model estimates into probability units. This allowed for a more direct and intuitive comparison against the effect sizes obtained from the standard LMMs used in our main analyses (estimates and confidence interval bounds for sleep efficiency were multiplied by 100 to match the original percentage scale).

Across all outcomes, effect estimates and their significance were highly consistent between the GLMM and LMM approaches (**Supplementary Tables 1-6**). The positive D1-N1 association between O3 and total sleep time, which was significant both before and after FDR correction in single-pollutant GLMM models, but not in LMM models, was only meaningful exception to such pattern. This association is, however, sensitive to FDR correction in multi-pollutant models, regardless of the approach. We therefore conclude that the results obtained from our original linear model analyses are robust and can be interpreted as reported.

### **Supplementary Figures and Tables**

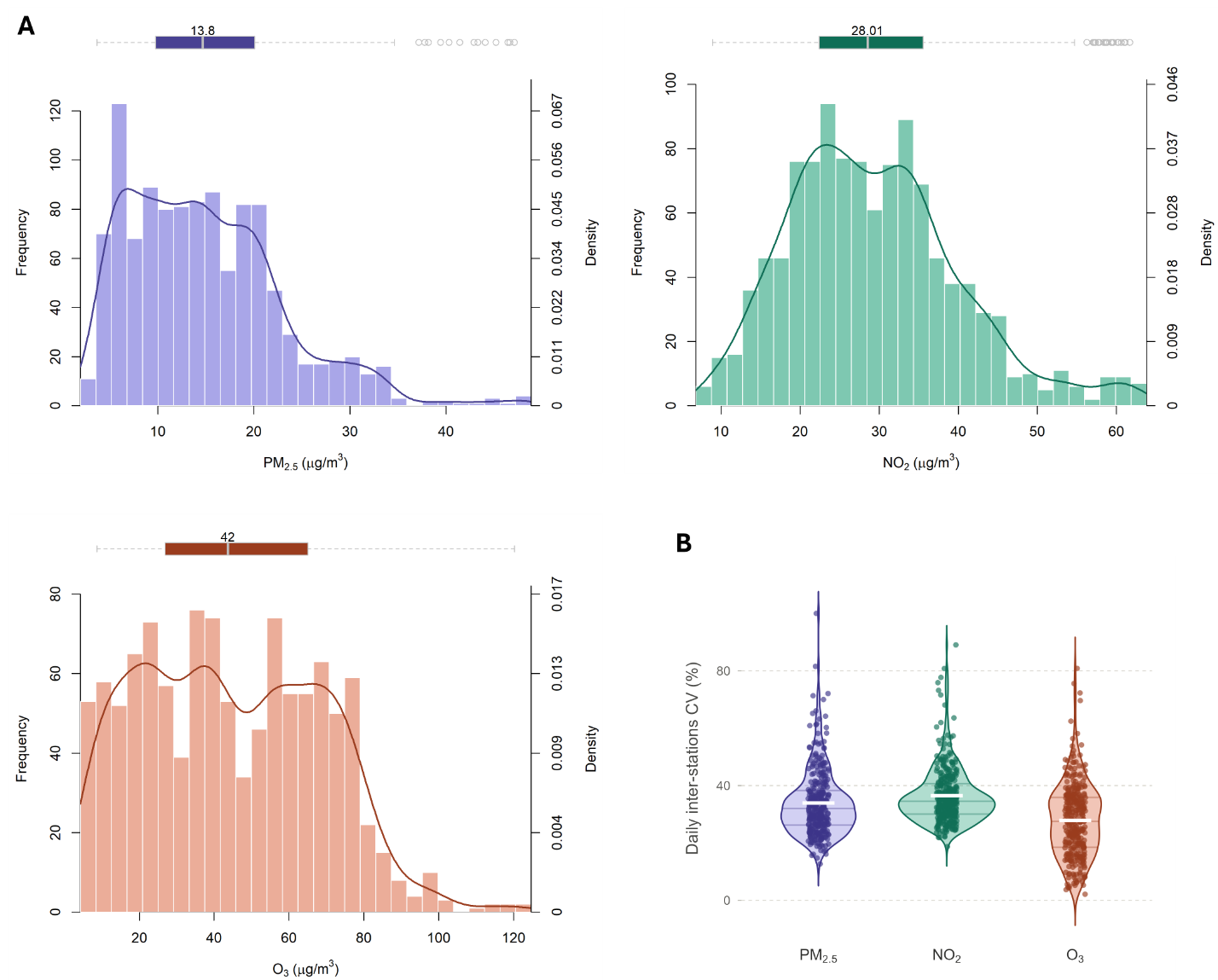

##### **Supplementary Figure 1.** Distribution of air pollutant concentrations.

(A) Distribution of pollutant concentrations at the observation level, illustrated with a histogram and superimposed density curve (dual y-axes) below, and a horizontal boxplot summarizing quartiles and outliers above. (B) Daily coefficient of variation (CV) for air pollutant concentrations across monitoring stations.

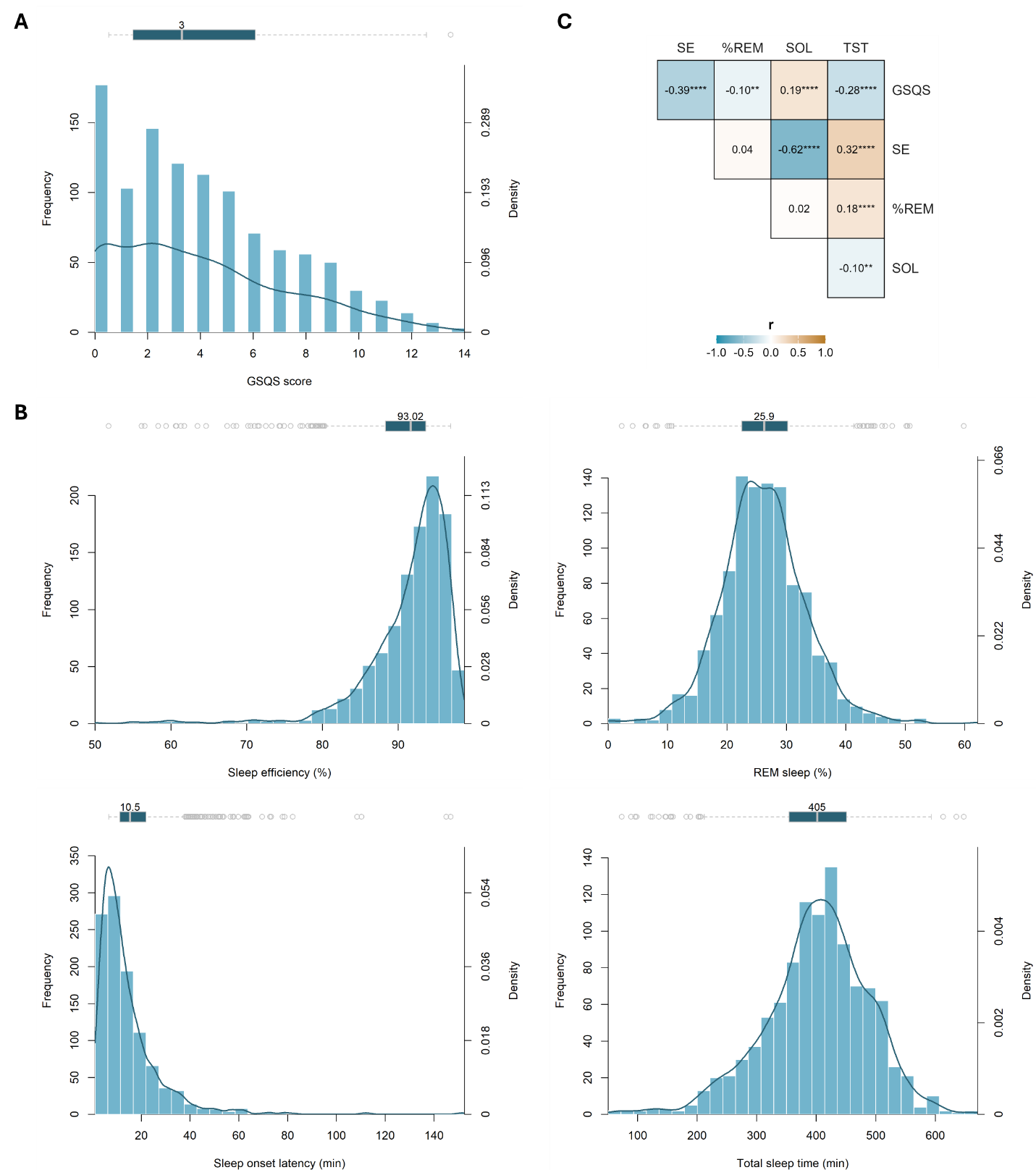

##### **Supplementary Figure 2.** Distribution and inter-correlations of sleep outcomes.

(**A-B**) The distribution of self-reported (Groningen Sleep Questionnaire Scale (GSQS); **A**) and device-measured (sleep efficiency (SE), % REM sleep, sleep onset latency (SOL), total sleep time (TST); **B**) sleep metrics is illustrated at the observation-level with a histogram and superimposed density curve (dual y-axes) below, and a horizontal boxplot summarizing quartiles and outliers above. (**C**) Pearson’s correlations across the abovementioned sleep metrics, displaying *r* and *p* values (** *p* < 0.01, **** *p* < 0.0001).

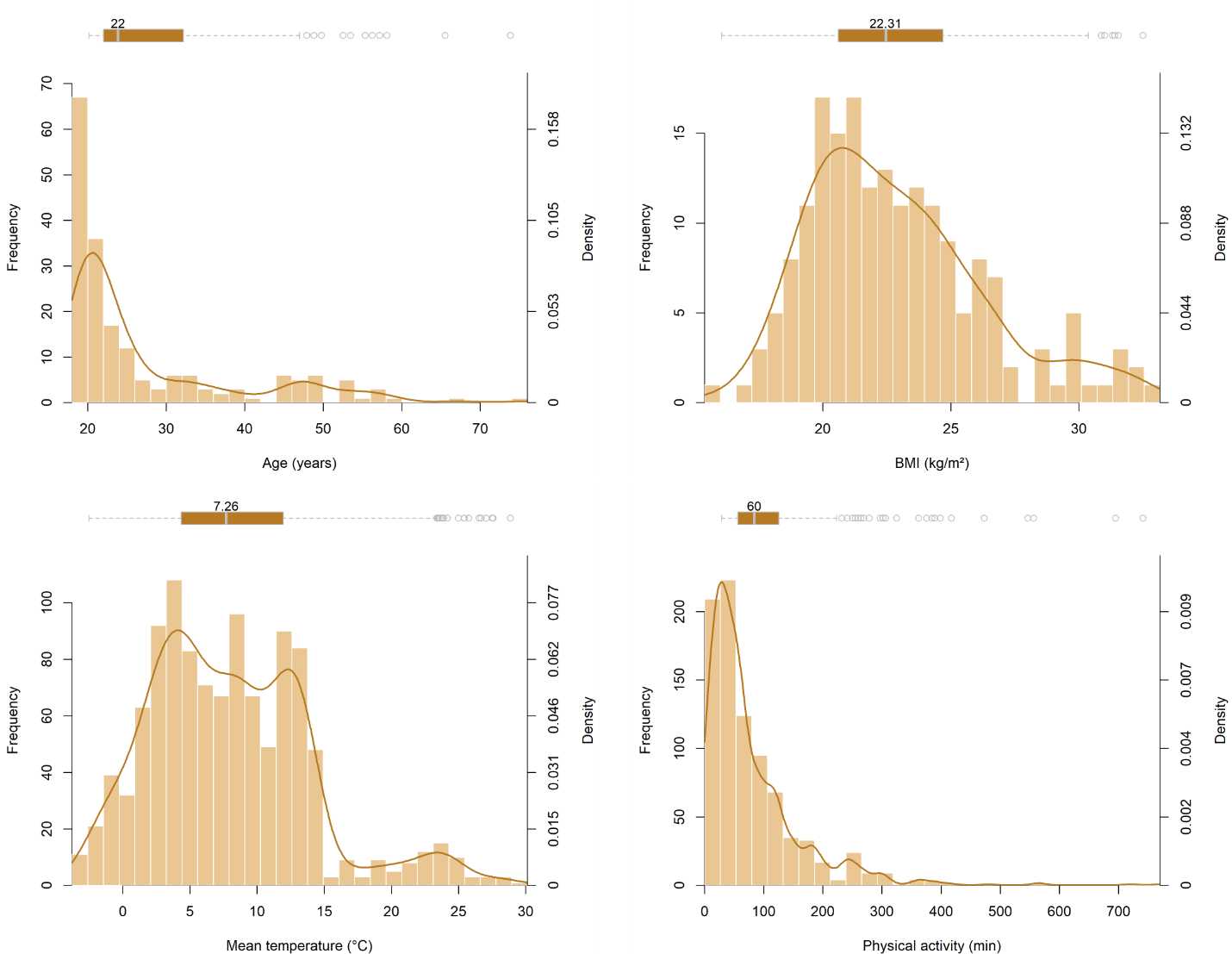

##### **Supplementary Figure 3.** Distribution of continuous covariates.

Distribution of static covariates at the subject-level (top) and dynamic covariates at the observation-level (bottom) is illustrated with a histogram and superimposed density curve (dual y-axes) below, and a horizontal boxplot summarizing quartiles and outliers above.

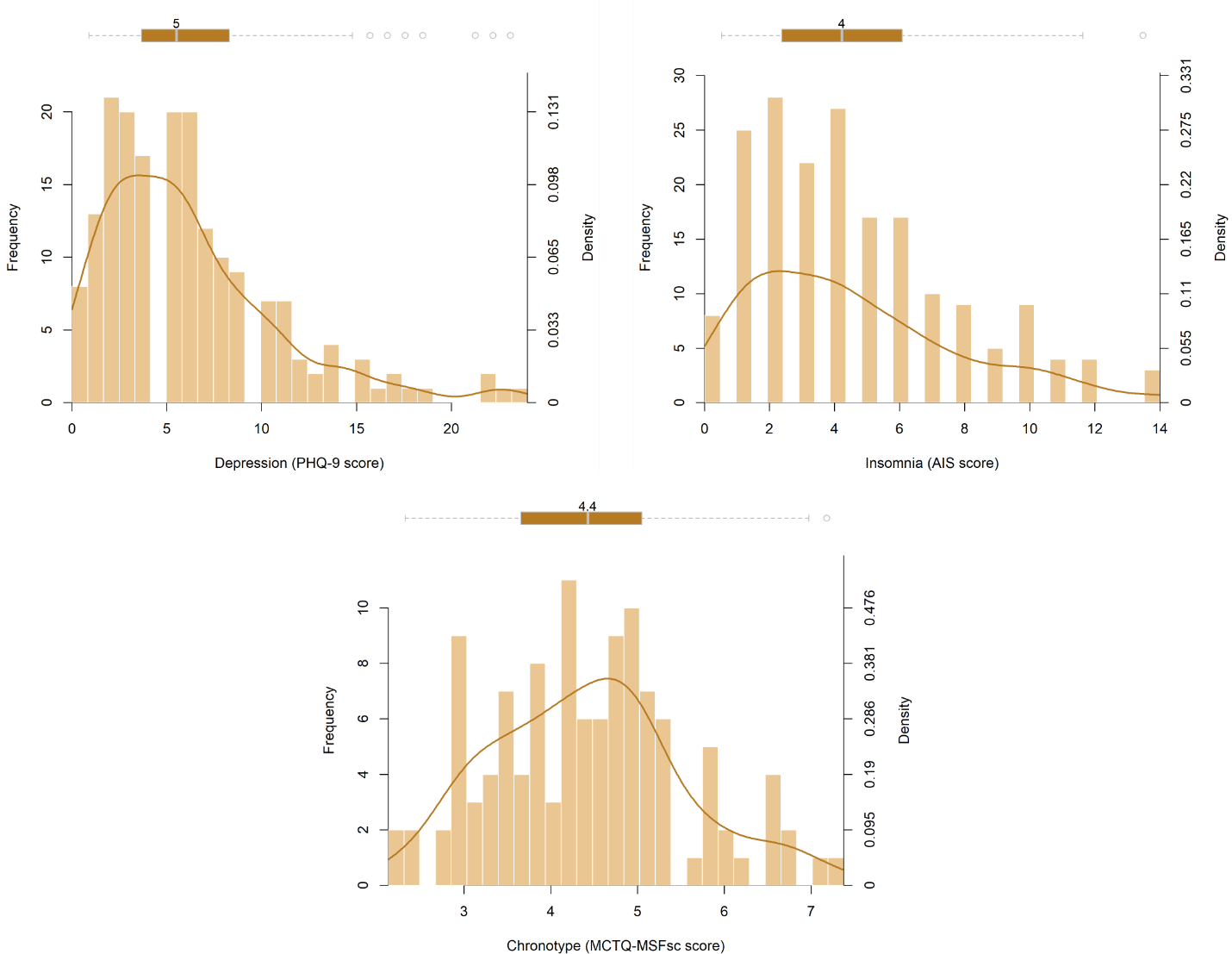

##### **Supplementary Figure 4.** Distribution of psychological and sleep-related measures used in sensitivity analyses.

Variable distribution is illustrated at the subject-level with a histogram and superimposed density curve (dual y-axes) below, and a horizontal boxplot summarizing quartiles and outliers above.

##### **Supplementary Table 1.** Statistical results for D1-N1 associations between PM_2.5_ and sleep outcomes.

| **PM_2.5_ (D1-N1)** | | **Single-Pollutant Model** | | | | **Multi-Pollutant Model** | | | |
| --- | --- | --- | --- | --- | --- | --- | --- | --- | --- |
|  | **Outcome** | **Estimate** | **SE** | **P-value** | **Q-value** | **Estimate** | **SE** | **P-value** | **Q-value** |
| **Main** | **GSQS** | 0.0001 | 0.0174 | 0.9959 | 0.9959 | 0.0062 | 0.0224 | 0.7815 | 0.9071 |
|  | **%SE** | 0.0124 | 0.0269 | 0.6452 | 0.9959 | 0.0054 | 0.0344 | 0.8750 | 0.9071 |
|  | **%REM** | 0.0070 | 0.0371 | 0.8502 | 0.9959 | -0.0122 | 0.0474 | 0.7970 | 0.9071 |
|  | **SOL** | -0.0326 | 0.0685 | 0.6339 | 0.9959 | -0.0513 | 0.0873 | 0.5569 | 0.9071 |
|  | **TST** | -0.0550 | 0.4623 | 0.9054 | 0.9959 | -0.0686 | 0.5878 | 0.9071 | 0.9071 |
| S1: Depression | **GSQS** | -0.0005 | 0.0172 | 0.9761 | 0.9761 | 0.0077 | 0.0221 | 0.7278 | 0.9337 |
|  | **%SE** | 0.0128 | 0.0271 | 0.6369 | 0.9761 | 0.0045 | 0.0346 | 0.8959 | 0.9337 |
|  | **%REM** | 0.0065 | 0.0374 | 0.8616 | 0.9761 | -0.0114 | 0.0478 | 0.8114 | 0.9337 |
|  | **SOL** | -0.0320 | 0.0691 | 0.6438 | 0.9761 | -0.0503 | 0.0881 | 0.5683 | 0.9337 |
|  | **TST** | -0.0361 | 0.4646 | 0.9381 | 0.9761 | -0.0492 | 0.5907 | 0.9337 | 0.9337 |
| S2: Insomnia | **GSQS** | -0.0045 | 0.0170 | 0.7904 | 0.9955 | 0.0023 | 0.0218 | 0.9149 | 0.9960 |
|  | **%SE** | 0.0137 | 0.0269 | 0.6103 | 0.9955 | 0.0066 | 0.0344 | 0.8480 | 0.9960 |
|  | **%REM** | 0.0063 | 0.0372 | 0.8655 | 0.9955 | -0.0128 | 0.0475 | 0.7877 | 0.9960 |
|  | **SOL** | -0.0344 | 0.0687 | 0.6169 | 0.9955 | -0.0534 | 0.0876 | 0.5421 | 0.9960 |
|  | **TST** | -0.0026 | 0.4611 | 0.9955 | 0.9955 | 0.0030 | 0.5861 | 0.9960 | 0.9960 |
| S3: Chronotype | **GSQS** | -0.0188 | 0.0225 | 0.4026 | 0.4026 | -0.0156 | 0.0284 | 0.5833 | 0.7291 |
|  | **%SE** | 0.0771 | 0.0337 | **0.0225** | 0.1125 | 0.0955 | 0.0422 | **0.0242** | 0.1210 |
|  | **%REM** | 0.0458 | 0.0482 | 0.3422 | 0.4026 | -0.0087 | 0.0602 | 0.8854 | 0.8854 |
|  | **SOL** | -0.0953 | 0.0926 | 0.3042 | 0.4026 | -0.1726 | 0.1160 | 0.1373 | 0.3433 |
|  | **TST** | 0.5763 | 0.5706 | 0.3130 | 0.4026 | 0.4402 | 0.7138 | 0.5378 | 0.7291 |
| S4: N0 Sleep | **GSQS** | -0.0190 | 0.0214 | 0.3747 | 0.4648 | -0.0164 | 0.0260 | 0.5303 | 0.6075 |
|  | **%SE** | 0.0384 | 0.0306 | 0.2105 | 0.4648 | 0.0359 | 0.0367 | 0.3290 | 0.5483 |
|  | **%REM** | 0.0315 | 0.0430 | 0.4648 | 0.4648 | 0.0265 | 0.0516 | 0.6075 | 0.6075 |
|  | **SOL** | -0.0989 | 0.0760 | 0.1934 | 0.4648 | -0.1130 | 0.0906 | 0.2131 | 0.5483 |
|  | **TST** | 0.5873 | 0.5557 | 0.2910 | 0.4648 | 0.6563 | 0.6630 | 0.3227 | 0.5483 |
| S5: GLMM | **GSQS** | 0.0003 | 0.0173 | 0.9855 | 0.9855 | 0.0068 | 0.0221 | 0.7577 | 0.9537 |
|  | **%SE** | 0.0118 | 0.0223 | 0.5963 | 0.9855 | 0.0037 | 0.0282 | 0.8946 | 0.9537 |
|  | **%REM** | 0.0070 | 0.0368 | 0.8489 | 0.9855 | -0.0112 | 0.0469 | 0.8106 | 0.9537 |
|  | **SOL** | -0.0294 | 0.0512 | 0.5657 | 0.9855 | -0.0553 | 0.0658 | 0.4008 | 0.9537 |
|  | **TST** | -0.0353 | 0.4590 | 0.9386 | 0.9855 | -0.0339 | 0.5842 | 0.9537 | 0.9537 |

##### **Supplementary Table 2.** Statistical results for D1-N1 associations between NO_2_ and sleep outcomes.

| **NO_2_ (D1-N1)** | | **Single-Pollutant Model** | | | | **Multi-Pollutant Model** | | | |
| --- | --- | --- | --- | --- | --- | --- | --- | --- | --- |
|  | **Outcome** | **Estimate** | **SE** | **P-value** | **Q-value** | **Estimate** | **SE** | **P-value** | **Q-value** |
| **Main** | **GSQS** | -0.0011 | 0.0126 | 0.9328 | 0.9328 | 0.0004 | 0.0158 | 0.9797 | 0.9955 |
|  | **%SE** | 0.0070 | 0.0195 | 0.7195 | 0.8994 | 0.0001 | 0.0245 | 0.9955 | 0.9955 |
|  | **%REM** | 0.0137 | 0.0269 | 0.6109 | 0.8994 | 0.0135 | 0.0339 | 0.6898 | 0.9955 |
|  | **SOL** | 0.0179 | 0.0496 | 0.7180 | 0.8994 | 0.0599 | 0.0623 | 0.3367 | 0.8418 |
|  | **TST** | 0.2121 | 0.3341 | 0.5258 | 0.8994 | 0.4897 | 0.4188 | 0.2427 | 0.8418 |
| S1: Depression | **GSQS** | -0.0038 | 0.0124 | 0.7585 | 0.7585 | -0.0040 | 0.0157 | 0.7984 | 0.9448 |
|  | **%SE** | 0.0083 | 0.0196 | 0.6735 | 0.7585 | 0.0017 | 0.0248 | 0.9448 | 0.9448 |
|  | **%REM** | 0.0120 | 0.0271 | 0.6572 | 0.7585 | 0.0109 | 0.0343 | 0.7515 | 0.9448 |
|  | **SOL** | 0.0180 | 0.0501 | 0.7197 | 0.7585 | 0.0607 | 0.0631 | 0.3365 | 0.8413 |
|  | **TST** | 0.2347 | 0.3363 | 0.4855 | 0.7585 | 0.5308 | 0.4223 | 0.2093 | 0.8413 |
| S2: Insomnia | **GSQS** | -0.0043 | 0.0122 | 0.7257 | 0.7279 | -0.0021 | 0.0154 | 0.8890 | 0.9706 |
|  | **%SE** | 0.0080 | 0.0195 | 0.6832 | 0.7279 | 0.0009 | 0.0246 | 0.9706 | 0.9706 |
|  | **%REM** | 0.0128 | 0.0269 | 0.6341 | 0.7279 | 0.0123 | 0.0339 | 0.7164 | 0.9706 |
|  | **SOL** | 0.0173 | 0.0497 | 0.7279 | 0.7279 | 0.0602 | 0.0625 | 0.3363 | 0.8408 |
|  | **TST** | 0.2342 | 0.3333 | 0.4824 | 0.7279 | 0.4945 | 0.4175 | 0.2366 | 0.8408 |
| S3: Chronotype | **GSQS** | -0.0069 | 0.0162 | 0.6696 | 0.6900 | 0.0038 | 0.0203 | 0.8510 | 0.8510 |
|  | **%SE** | 0.0160 | 0.0242 | 0.5081 | 0.6900 | -0.0246 | 0.0300 | 0.4131 | 0.5164 |
|  | **%REM** | 0.0612 | 0.0343 | 0.0751 | 0.3755 | 0.0617 | 0.0428 | 0.1500 | 0.5164 |
|  | **SOL** | 0.0265 | 0.0663 | 0.6900 | 0.6900 | 0.1003 | 0.0822 | 0.2233 | 0.5164 |
|  | **TST** | 0.4800 | 0.4074 | 0.2393 | 0.5983 | 0.4549 | 0.5065 | 0.3696 | 0.5164 |
| S4: N0 Sleep | **GSQS** | -0.0079 | 0.0149 | 0.5980 | 0.8268 | 0.0000 | 0.0180 | 0.9986 | 0.9986 |
|  | **%SE** | 0.0131 | 0.0216 | 0.5441 | 0.8268 | -0.0035 | 0.0256 | 0.8902 | 0.9986 |
|  | **%REM** | 0.0133 | 0.0304 | 0.6614 | 0.8268 | 0.0006 | 0.0361 | 0.9867 | 0.9986 |
|  | **SOL** | -0.0079 | 0.0532 | 0.8821 | 0.8821 | 0.0460 | 0.0630 | 0.4651 | 0.9986 |
|  | **TST** | 0.3732 | 0.3898 | 0.3389 | 0.8268 | 0.3405 | 0.4617 | 0.4611 | 0.9986 |
| S5: GLMM | **GSQS** | -0.0012 | 0.0124 | 0.9211 | 0.9211 | -0.0001 | 0.0157 | 0.9955 | 0.9955 |
|  | **%SE** | 0.0076 | 0.0160 | 0.6355 | 0.8308 | 0.0015 | 0.0202 | 0.9426 | 0.9955 |
|  | **%REM** | 0.0130 | 0.0267 | 0.6258 | 0.8308 | 0.0126 | 0.0335 | 0.7066 | 0.9955 |
|  | **SOL** | 0.0162 | 0.0373 | 0.6646 | 0.8308 | 0.0517 | 0.0474 | 0.2747 | 0.6868 |
|  | **TST** | 0.2141 | 0.3307 | 0.5174 | 0.8308 | 0.4765 | 0.4142 | 0.2500 | 0.6868 |

##### **Supplementary Table 3.** Statistical results for D1-N1 associations between O_3_ and sleep outcomes.

| **O_3_ (D1-N1)** | | **Single-Pollutant Model** | | | | **Multi-Pollutant Model** | | | |
| --- | --- | --- | --- | --- | --- | --- | --- | --- | --- |
|  | **Outcome** | **Estimate** | **SE** | **P-value** | **Q-value** | **Estimate** | **SE** | **P-value** | **Q-value** |
| **Main** | **GSQS** | 0.0082 | 0.0094 | 0.3872 | 0.5270 | 0.0095 | 0.0103 | 0.3553 | 0.5922 |
|  | **%SE** | -0.0114 | 0.0149 | 0.4453 | 0.5270 | -0.0102 | 0.0163 | 0.5304 | 0.5946 |
|  | **%REM** | -0.0131 | 0.0206 | 0.5270 | 0.5270 | -0.0120 | 0.0226 | 0.5946 | 0.5946 |
|  | **SOL** | 0.0403 | 0.0380 | 0.2887 | 0.5270 | 0.0452 | 0.0414 | 0.2755 | 0.5922 |
|  | **TST** | 0.4748 | 0.2541 | 0.0621 | 0.3105 | 0.5820 | 0.2756 | **0.0351** | 0.1755 |
| S1: Depression | **GSQS** | 0.0068 | 0.0093 | 0.4642 | 0.5054 | 0.0074 | 0.0101 | 0.4667 | 0.5594 |
|  | **%SE** | -0.0117 | 0.0151 | 0.4401 | 0.5054 | -0.0103 | 0.0165 | 0.5347 | 0.5594 |
|  | **%REM** | -0.0139 | 0.0209 | 0.5054 | 0.5054 | -0.0134 | 0.0229 | 0.5594 | 0.5594 |
|  | **SOL** | 0.0412 | 0.0385 | 0.2842 | 0.5054 | 0.0469 | 0.0421 | 0.2657 | 0.5594 |
|  | **TST** | 0.5146 | 0.2565 | **0.0453** | 0.2265 | 0.6382 | 0.2781 | **0.0221** | 0.1105 |
| S2: Insomnia | **GSQS** | 0.0077 | 0.0091 | 0.3967 | 0.5031 | 0.0077 | 0.0098 | 0.4347 | 0.5599 |
|  | **%SE** | -0.0111 | 0.0149 | 0.4566 | 0.5031 | -0.0095 | 0.0163 | 0.5599 | 0.5599 |
|  | **%REM** | -0.0138 | 0.0207 | 0.5031 | 0.5031 | -0.0133 | 0.0227 | 0.5589 | 0.5599 |
|  | **SOL** | 0.0402 | 0.0380 | 0.2906 | 0.5031 | 0.0448 | 0.0416 | 0.2812 | 0.5599 |
|  | **TST** | 0.4931 | 0.2530 | 0.0517 | 0.2585 | 0.6152 | 0.2740 | **0.0251** | 0.1255 |
| S3: Chronotype | **GSQS** | 0.0124 | 0.0124 | 0.3186 | 0.6166 | 0.0104 | 0.0134 | 0.4370 | 0.8978 |
|  | **%SE** | -0.0144 | 0.0189 | 0.4465 | 0.6166 | -0.0026 | 0.0203 | 0.8963 | 0.8978 |
|  | **%REM** | -0.0215 | 0.0270 | 0.4251 | 0.6166 | -0.0073 | 0.0291 | 0.8014 | 0.8978 |
|  | **SOL** | 0.0144 | 0.0510 | 0.7783 | 0.7783 | 0.0070 | 0.0543 | 0.8978 | 0.8978 |
|  | **TST** | 0.2171 | 0.3167 | 0.4933 | 0.6166 | 0.4050 | 0.3388 | 0.2328 | 0.8978 |
| S4: N0 Sleep | **GSQS** | 0.0069 | 0.0107 | 0.5207 | 0.5785 | 0.0044 | 0.0113 | 0.6985 | 0.7165 |
|  | **%SE** | -0.0141 | 0.0159 | 0.3769 | 0.5785 | -0.0094 | 0.0167 | 0.5728 | 0.7165 |
|  | **%REM** | -0.0123 | 0.0222 | 0.5785 | 0.5785 | -0.0085 | 0.0233 | 0.7165 | 0.7165 |
|  | **SOL** | 0.0407 | 0.0397 | 0.3050 | 0.5785 | 0.0323 | 0.0426 | 0.4487 | 0.7165 |
|  | **TST** | 0.3844 | 0.2894 | 0.1846 | 0.5785 | 0.5584 | 0.3067 | 0.0693 | 0.3465 |
| S5: GLMM | **GSQS** | 0.0082 | 0.0093 | 0.3764 | 0.5315 | 0.0096 | 0.0101 | 0.3437 | 0.5478 |
|  | **%SE** | -0.0116 | 0.0124 | 0.3475 | 0.5315 | -0.0105 | 0.0135 | 0.4359 | 0.5478 |
|  | **%REM** | -0.0128 | 0.0204 | 0.5315 | 0.5315 | -0.0118 | 0.0222 | 0.5952 | 0.5952 |
|  | **SOL** | 0.0228 | 0.0295 | 0.4387 | 0.5315 | 0.0251 | 0.0324 | 0.4382 | 0.5478 |
|  | **TST** | 0.6225 | 0.2403 | **0.0096** | **0.0480** | 0.5952 | 0.2714 | **0.0283** | 0.1415 |

##### **Supplementary Table 4.** Statistical results for D1-N2 associations between PM_2.5_ and sleep outcomes.

| **PM_2.5_ (D1-N2)** | | **Single-Pollutant Model** | | | | **Multi-Pollutant Model** | | | |
| --- | --- | --- | --- | --- | --- | --- | --- | --- | --- |
|  | **Outcome** | **Estimate** | **SE** | **P-value** | **Q-value** | **Estimate** | **SE** | **P-value** | **Q-value** |
| **Main** | **GSQS** | -0.0037 | 0.0169 | 0.8249 | 0.8249 | 0.0554 | 0.0215 | **0.0103** | 0.0515 |
|  | **%SE** | 0.0386 | 0.0259 | 0.1370 | 0.3425 | -0.0322 | 0.0331 | 0.3314 | 0.4142 |
|  | **%REM** | 0.0763 | 0.0360 | **0.0345** | 0.1725 | 0.0813 | 0.0465 | 0.0806 | 0.2015 |
|  | **SOL** | -0.0172 | 0.0608 | 0.7773 | 0.8249 | 0.0855 | 0.0782 | 0.2748 | 0.4142 |
|  | **TST** | 0.3806 | 0.4449 | 0.3926 | 0.6543 | 0.1720 | 0.5754 | 0.7651 | 0.7651 |
| S1: Depression | **GSQS** | -0.0066 | 0.0168 | 0.6967 | 0.8191 | 0.0547 | 0.0214 | **0.0109** | 0.0545 |
|  | **%SE** | 0.0393 | 0.0263 | 0.1353 | 0.3383 | -0.0337 | 0.0335 | 0.3159 | 0.3949 |
|  | **%REM** | 0.0733 | 0.0366 | **0.0454** | 0.2270 | 0.0780 | 0.0471 | 0.0986 | 0.2465 |
|  | **SOL** | -0.0141 | 0.0617 | 0.8191 | 0.8191 | 0.0941 | 0.0794 | 0.2360 | 0.3933 |
|  | **TST** | 0.4246 | 0.4505 | 0.3462 | 0.5770 | 0.1999 | 0.5823 | 0.7315 | 0.7315 |
| S2: Insomnia | **GSQS** | -0.0083 | 0.0166 | 0.6160 | 0.7427 | 0.0478 | 0.0212 | **0.0243** | 0.1215 |
|  | **%SE** | 0.0407 | 0.0260 | 0.1176 | 0.2940 | -0.0288 | 0.0332 | 0.3863 | 0.4829 |
|  | **%REM** | 0.0741 | 0.0362 | **0.0410** | 0.2050 | 0.0770 | 0.0467 | 0.0996 | 0.2490 |
|  | **SOL** | -0.0201 | 0.0611 | 0.7427 | 0.7427 | 0.0818 | 0.0787 | 0.2988 | 0.4829 |
|  | **TST** | 0.4305 | 0.4454 | 0.3341 | 0.5568 | 0.2619 | 0.5764 | 0.6497 | 0.6497 |
| S3: Chronotype | **GSQS** | -0.0027 | 0.0224 | 0.9049 | 0.9049 | 0.0664 | 0.0274 | **0.0156** | 0.0780 |
|  | **%SE** | 0.0552 | 0.0324 | 0.0892 | 0.3185 | -0.0226 | 0.0397 | 0.5699 | 0.7254 |
|  | **%REM** | 0.0720 | 0.0471 | 0.1274 | 0.3185 | 0.0412 | 0.0586 | 0.4827 | 0.7254 |
|  | **SOL** | -0.0358 | 0.0789 | 0.6499 | 0.8124 | 0.0542 | 0.0980 | 0.5803 | 0.7254 |
|  | **TST** | 0.3424 | 0.5638 | 0.5439 | 0.8124 | -0.0025 | 0.7051 | 0.9972 | 0.9972 |
| S4: N1 Sleep | **GSQS** | -0.0135 | 0.0192 | 0.4845 | 0.8075 | 0.0366 | 0.0245 | 0.1351 | 0.6755 |
|  | **%SE** | 0.0373 | 0.0273 | 0.1724 | 0.6865 | -0.0170 | 0.0346 | 0.6237 | 0.7796 |
|  | **%REM** | 0.0421 | 0.0385 | 0.2746 | 0.6865 | 0.0312 | 0.0490 | 0.5253 | 0.7796 |
|  | **SOL** | 0.0017 | 0.0673 | 0.9800 | 0.9800 | 0.0543 | 0.0858 | 0.5269 | 0.7796 |
|  | **TST** | 0.1955 | 0.4942 | 0.6926 | 0.8658 | -0.1660 | 0.6295 | 0.7921 | 0.7921 |
| S5: GLMM | **GSQS** | -0.0038 | 0.0167 | 0.8208 | 0.8208 | 0.0550 | 0.0213 | **0.0098** | **0.0490** |
|  | **%SE** | 0.0370 | 0.0213 | 0.0814 | 0.2035 | -0.0208 | 0.0270 | 0.4415 | 0.4415 |
|  | **%REM** | 0.0773 | 0.0357 | **0.0306** | 0.1530 | 0.0827 | 0.0461 | 0.0726 | 0.1815 |
|  | **SOL** | -0.0191 | 0.0506 | 0.7050 | 0.8208 | 0.0766 | 0.0644 | 0.2344 | 0.3248 |
|  | **TST** | 0.3970 | 0.4410 | 0.3680 | 0.6133 | 0.6396 | 0.5676 | 0.2598 | 0.3248 |
| S6: Awake time | **GSQS** | -0.0050 | 0.0186 | 0.7903 | 0.7903 | 0.0393 | 0.0235 | 0.0943 | 0.4715 |
|  | **%SE** | 0.0420 | 0.0268 | 0.1176 | 0.2940 | -0.0197 | 0.0337 | 0.5587 | 0.7418 |
|  | **%REM** | 0.0646 | 0.0379 | 0.0887 | 0.2940 | 0.0522 | 0.0481 | 0.2779 | 0.6948 |
|  | **SOL** | -0.0245 | 0.0659 | 0.7100 | 0.7903 | 0.0448 | 0.0838 | 0.5934 | 0.7418 |
|  | **TST** | 0.1521 | 0.4371 | 0.7280 | 0.7903 | -0.0962 | 0.5544 | 0.8623 | 0.8623 |

##### **Supplementary Table 5.** Statistical results for D1-N2 associations between NO_2_ and sleep outcomes.

| **NO_2_ (D1-N2)** | | **Single-Pollutant Model** | | | | **Multi-Pollutant Model** | | | |
| --- | --- | --- | --- | --- | --- | --- | --- | --- | --- |
|  | **Outcome** | **Estimate** | **SE** | **P-value** | **Q-value** | **Estimate** | **SE** | **P-value** | **Q-value** |
| **Main** | **GSQS** | -0.0306 | 0.0115 | **0.0081** | **0.0202** | -0.0329 | 0.0149 | **0.0276** | 0.1380 |
|  | **%SE** | 0.0489 | 0.0177 | **0.0059** | **0.0202** | 0.0312 | 0.0230 | 0.1740 | 0.4350 |
|  | **%REM** | 0.0382 | 0.0247 | 0.1227 | 0.2045 | 0.0226 | 0.0323 | 0.4845 | 0.6056 |
|  | **SOL** | -0.0547 | 0.0416 | 0.1891 | 0.2364 | -0.0443 | 0.0543 | 0.4154 | 0.6056 |
|  | **TST** | 0.2815 | 0.3051 | 0.3566 | 0.3566 | 0.1710 | 0.3980 | 0.6675 | 0.6675 |
| S1: Depression | **GSQS** | -0.0337 | 0.0114 | **0.0033** | **0.0123** | -0.0368 | 0.0148 | **0.0129** | 0.0645 |
|  | **%SE** | 0.0505 | 0.0179 | **0.0049** | **0.0123** | 0.0330 | 0.0232 | 0.1560 | 0.3900 |
|  | **%REM** | 0.0365 | 0.0250 | 0.1448 | 0.2205 | 0.0216 | 0.0327 | 0.5092 | 0.6229 |
|  | **SOL** | -0.0569 | 0.0421 | 0.1764 | 0.2205 | -0.0492 | 0.0550 | 0.3718 | 0.6197 |
|  | **TST** | 0.3138 | 0.3079 | 0.3086 | 0.3086 | 0.1978 | 0.4020 | 0.6229 | 0.6229 |
| S2: Insomnia | **GSQS** | -0.0311 | 0.0113 | **0.0061** | **0.0153** | -0.0326 | 0.0146 | **0.0255** | 0.1275 |
|  | **%SE** | 0.0489 | 0.0177 | **0.0059** | **0.0153** | 0.0304 | 0.0230 | 0.1855 | 0.4638 |
|  | **%REM** | 0.0382 | 0.0248 | 0.1235 | 0.2058 | 0.0234 | 0.0323 | 0.4699 | 0.5874 |
|  | **SOL** | -0.0551 | 0.0417 | 0.1873 | 0.2341 | -0.0434 | 0.0545 | 0.4257 | 0.5874 |
|  | **TST** | 0.2844 | 0.3048 | 0.3511 | 0.3511 | 0.1549 | 0.3971 | 0.6966 | 0.6966 |
| S3: Chronotype | **GSQS** | -0.0419 | 0.0150 | **0.0053** | **0.0133** | -0.0469 | 0.0187 | **0.0123** | 0.0615 |
|  | **%SE** | 0.0658 | 0.0215 | **0.0024** | **0.0120** | 0.0465 | 0.0271 | 0.0875 | 0.2188 |
|  | **%REM** | 0.0563 | 0.0316 | 0.0751 | 0.1252 | 0.0490 | 0.0401 | 0.2224 | 0.3707 |
|  | **SOL** | -0.0545 | 0.0529 | 0.3028 | 0.3232 | -0.0251 | 0.0669 | 0.7070 | 0.7070 |
|  | **TST** | 0.3737 | 0.3779 | 0.3232 | 0.3232 | 0.3458 | 0.4807 | 0.4723 | 0.5904 |
| S4: N1 Sleep | **GSQS** | -0.0352 | 0.0131 | **0.0075** | **0.0375** | -0.0395 | 0.0168 | **0.0190** | 0.0950 |
|  | **%SE** | 0.0438 | 0.0189 | **0.0206** | **0.0515** | 0.0300 | 0.0243 | 0.2163 | 0.5287 |
|  | **%REM** | 0.0332 | 0.0267 | 0.2129 | 0.3548 | 0.0343 | 0.0343 | 0.3172 | 0.5287 |
|  | **SOL** | -0.0296 | 0.0466 | 0.5258 | 0.5258 | -0.0353 | 0.0607 | 0.5613 | 0.5613 |
|  | **TST** | 0.2928 | 0.3422 | 0.3926 | 0.4908 | 0.2829 | 0.4432 | 0.5235 | 0.5613 |
| S5: GLMM | **GSQS** | -0.0305 | 0.0114 | **0.0076** | **0.0190** | -0.0329 | 0.0147 | **0.0252** | 0.1260 |
|  | **%SE** | 0.0428 | 0.0146 | **0.0034** | **0.0170** | 0.0254 | 0.0191 | 0.1822 | 0.3807 |
|  | **%REM** | 0.0380 | 0.0245 | 0.1202 | 0.1503 | 0.0212 | 0.0320 | 0.5078 | 0.6348 |
|  | **SOL** | -0.0589 | 0.0349 | 0.0913 | 0.1503 | -0.0540 | 0.0448 | 0.2284 | 0.3807 |
|  | **TST** | 0.2250 | 0.3041 | 0.4595 | 0.4595 | 0.0347 | 0.3914 | 0.9294 | 0.9294 |
| S6: Awake time | **GSQS** | -0.0261 | 0.0127 | **0.0405** | 0.1013 | -0.0275 | 0.0162 | 0.0910 | 0.3580 |
|  | **%SE** | 0.0484 | 0.0183 | **0.0083** | **0.0415** | 0.0306 | 0.0235 | 0.1938 | 0.3580 |
|  | **%REM** | 0.0451 | 0.0259 | 0.0827 | 0.1378 | 0.0416 | 0.0335 | 0.2148 | 0.3580 |
|  | **SOL** | -0.0466 | 0.0451 | 0.3020 | 0.3775 | -0.0373 | 0.0587 | 0.5253 | 0.5253 |
|  | **TST** | 0.2320 | 0.3002 | 0.4400 | 0.4400 | 0.2606 | 0.3830 | 0.4965 | 0.5253 |

##### **Supplementary Table 6.** Statistical results for D1-N2 associations between O_3_ and sleep outcomes.

| **O_3_ (D1-N2)** | | **Single-Pollutant Model** | | | | **Multi-Pollutant Model** | | | |
| --- | --- | --- | --- | --- | --- | --- | --- | --- | --- |
|  | **Outcome** | **Estimate** | **SE** | **P-value** | **Q-value** | **Estimate** | **SE** | **P-value** | **Q-value** |
| **Main** | **GSQS** | 0.0373 | 0.0093 | **0.0001** | **0.0003** | 0.0388 | 0.0107 | **0.0003** | **0.0015** |
|  | **%SE** | -0.0602 | 0.0145 | **0.0000** | **0.0000** | -0.0563 | 0.0167 | **0.0008** | **0.0020** |
|  | **%REM** | 0.0040 | 0.0205 | 0.8459 | 0.8459 | 0.0327 | 0.0236 | 0.1664 | 0.2080 |
|  | **SOL** | 0.0780 | 0.0344 | **0.0236** | **0.0393** | 0.0823 | 0.0398 | **0.0391** | 0.0652 |
|  | **TST** | -0.1772 | 0.2503 | 0.4791 | 0.5989 | -0.0723 | 0.2857 | 0.8003 | 0.8003 |
| S1: Depression | **GSQS** | 0.0375 | 0.0093 | **0.0001** | **0.0003** | 0.0374 | 0.0105 | **0.0004** | **0.0018** |
|  | **%SE** | -0.0617 | 0.0147 | **0.0000** | **0.0000** | -0.0574 | 0.0169 | **0.0007** | **0.0018** |
|  | **%REM** | 0.0040 | 0.0208 | 0.8458 | 0.8458 | 0.0315 | 0.0240 | 0.1887 | 0.2359 |
|  | **SOL** | 0.0798 | 0.0349 | **0.0227** | **0.0378** | 0.0841 | 0.0404 | **0.0376** | 0.0627 |
|  | **TST** | -0.1836 | 0.2538 | 0.4696 | 0.5870 | -0.0616 | 0.2892 | 0.8315 | 0.8315 |
| S2: Insomnia | **GSQS** | 0.0353 | 0.0091 | **0.0001** | **0.0003** | 0.0354 | 0.0103 | **0.0006** | **0.0025** |
|  | **%SE** | -0.0597 | 0.0145 | **0.0000** | **0.0000** | -0.0554 | 0.0167 | **0.0010** | **0.0025** |
|  | **%REM** | 0.0028 | 0.0206 | 0.8908 | 0.8908 | 0.0308 | 0.0237 | 0.1936 | 0.2420 |
|  | **SOL** | 0.0781 | 0.0345 | **0.0241** | **0.0402** | 0.0818 | 0.0400 | **0.0411** | 0.0685 |
|  | **TST** | -0.1596 | 0.2501 | 0.5237 | 0.6546 | -0.0380 | 0.2852 | 0.8940 | 0.8940 |
| S3: Chronotype | **GSQS** | 0.0475 | 0.0125 | **0.0002** | **0.0005** | 0.0457 | 0.0137 | **0.0009** | **0.0045** |
|  | **%SE** | -0.0691 | 0.0182 | **0.0002** | **0.0005** | -0.0574 | 0.0203 | **0.0049** | **0.0123** |
|  | **%REM** | -0.0087 | 0.0269 | 0.7464 | 0.7464 | 0.0175 | 0.0300 | 0.5603 | 0.7004 |
|  | **SOL** | 0.1063 | 0.0445 | **0.0172** | **0.0287** | 0.1090 | 0.0495 | **0.0281** | **0.0468** |
|  | **TST** | -0.1780 | 0.3196 | 0.5779 | 0.7224 | -0.0612 | 0.3551 | 0.8633 | 0.8633 |
| S4: N1 Sleep | **GSQS** | 0.0261 | 0.0107 | **0.0148** | **0.0370** | 0.0211 | 0.0120 | 0.0787 | 0.1968 |
|  | **%SE** | -0.0473 | 0.0155 | **0.0024** | **0.0120** | -0.0407 | 0.0175 | **0.0208** | 0.1040 |
|  | **%REM** | 0.0070 | 0.0219 | 0.7498 | 0.7498 | 0.0261 | 0.0246 | 0.2896 | 0.4827 |
|  | **SOL** | 0.0306 | 0.0388 | 0.4315 | 0.5644 | 0.0303 | 0.0448 | 0.4994 | 0.6243 |
|  | **TST** | -0.2137 | 0.2836 | 0.4515 | 0.5644 | -0.1512 | 0.3230 | 0.6399 | 0.6399 |
| S5: GLMM | **GSQS** | 0.0370 | 0.0092 | **0.0001** | **0.0003** | 0.0385 | 0.0105 | **0.0002** | **0.0010** |
|  | **%SE** | -0.0498 | 0.0119 | **0.0000** | **0.0000** | -0.0449 | 0.0138 | **0.0012** | **0.0030** |
|  | **%REM** | 0.0036 | 0.0203 | 0.8585 | 0.8721 | 0.0319 | 0.0233 | 0.1709 | 0.2136 |
|  | **SOL** | 0.0681 | 0.0286 | **0.0170** | **0.0283** | 0.0666 | 0.0326 | **0.0408** | 0.0680 |
|  | **TST** | 0.0386 | 0.2398 | 0.8721 | 0.8721 | 0.1762 | 0.2619 | 0.5011 | 0.5011 |
| S6: Awake time | **GSQS** | 0.0295 | 0.0104 | **0.0047** | **0.0118** | 0.0289 | 0.0117 | **0.0135** | **0.0338** |
|  | **%SE** | -0.0551 | 0.0150 | **0.0003** | **0.0015** | -0.0485 | 0.0171 | **0.0048** | **0.0240** |
|  | **%REM** | 0.0046 | 0.0215 | 0.8324 | 0.8324 | 0.0325 | 0.0245 | 0.1854 | 0.3090 |
|  | **SOL** | 0.0527 | 0.0375 | 0.1599 | 0.2665 | 0.0491 | 0.0434 | 0.2578 | 0.3223 |
|  | **TST** | -0.0853 | 0.2451 | 0.7281 | 0.8324 | -0.0238 | 0.2718 | 0.9301 | 0.9301 |
